# Huntingtin CAG repeat is a continuous modifier of brain structure and health vulnerability

**DOI:** 10.64898/2026.05.08.26352223

**Authors:** Harriet Cullen, Christopher Clarkson, Henrique Nascimento, Matteo Zanovello, Jeffrey Long, Mark Caulfield, Michael Simpson, Sarah J Tabrizi, Arianna Tucci

## Abstract

Huntington’s disease is caused by a CAG repeat expansion in the Huntingtin gene (*HTT*) above a pathogenic threshold; however, the biological consequences of repeat-length variation below this threshold remain poorly understood. Using whole-genome sequencing and linked phenotypic data from UK Biobank participants, we show that repeat-length variation within the normal and intermediate range is associated with measurable differences in brain volume, neuropsychiatric risk, and cognitive processing, and that only one third of pathogenic allele carriers have a recorded clinical diagnosis.

Analyses were performed in 474,446 UK Biobank participants, including 30,052 with intermediate repeats (27*–*35), 873 with reduced-penetrance repeats (36*–*39), and 155 with pathogenic repeats (≥40); 48,378 individuals had structural MRI. For quantitative phenotypes (brain volumes and cognition), associations with continuous repeat length were modelled using linear regression within the normal and intermediate range (≤35 repeats); deviation at ≥36 repeats was defined as departure from the extrapolated linear trend. For clinical outcomes (depression, anxiety, dementia, and delirium), repeat length was analysed categorically using Kaplan–Meier and Cox proportional hazards models with age as the timescale.

Within the normal and intermediate range, longer *HTT* CAG repeat length was associated with smaller subcortical and global brain volumes, including the accumbens, putamen, thalamus, hippocampus, and total grey and white matter, with effects amplified in older individuals. Intermediate alleles were associated with an increase in age-dependent depression risk (HR = 1.05, 95% CI 1.02*–*1.10) and longer repeat length within the normal and intermediate range predicted faster reaction time, a pattern that reversed sharply at pathogenic lengths. Among carriers of 40–41 CAG repeats, only 42% (95% CI 19–59%) had received a recorded Huntington’s disease diagnosis by age 84; however, the majority of pathogenic allele carriers who underwent neuroimaging met biomarker criteria for Stage 1 disease, indicating that early neurodegeneration is present in these individuals.

This work challenges the current understanding of the *HTT* CAG repeat length as a purely categorical determinant of monogenic disease and shows that repeat length acts as a quantitative modifier of brain structure and neuropsychiatric vulnerability across the population. These findings have implications for risk prediction, penetrance estimation, and the interpretation of repeat variation in population genomics.

## 2. Introduction

As population cohorts grow, we are increasingly recognising that genetic risk is rarely binary. Variants once considered either causative or benign are now understood to act as quantitative modifiers of biological function^1^.

Huntington’s disease (HD) has long been the paradigmatic monogenic neurological disorder, caused by a CAG trinucleotide repeat expansion in the *HTT* gene^2^ with near-complete penetrance above 40 repeats. *HTT* CAG repeat length is interpreted using discrete categories of increasing repeat size—normal, intermediate, reduced-penetrance, and pathogenic. This categorical framework, whilst clinically useful, is biologically reductive, assuming that repeat lengths below pathogenic thresholds are biologically neutral. Here, using one of the largest population cohorts with linked genomic, neuroimaging and clinical data we interrogate this assumption directly.

HD is characterised by progressive motor, cognitive and psychiatric symptoms and by neurodegeneration that is particularly prominent in the striatum. HD shows substantial clinical heterogeneity, with repeat length influencing both age at onset and disease severity; longer expansions are generally associated with earlier and more severe disease^3^. Current understanding of the phenotypic consequences of increasing repeat length has largely been derived from clinically ascertained individuals and their family members. Much less is known about the effects of repeat-length variation in unselected population cohorts. Emerging evidence suggests that intermediate alleles, although not associated with classical monogenic disease, may nonetheless confer elevated risk for neurological, cognitive and psychiatric phenotypes^4,5^. These observations raise the possibility that germline *HTT* CAG repeat length may act as a quantitative modifier of brain biology and health vulnerability across the population, rather than solely as a categorical determinant of monogenic disease.

Historically, comprehensive assessment of repeat-length variation has been limited by technical constraints. This is changing with the emergence of large-scale population genomic datasets. Short-read whole-genome sequencing (sr-WGS) can now accurately quantify repeat lengths^6^, enabling systematic investigation of their phenotypic consequences across the full allelic spectrum. In a previous population-scale analysis in the 100,000 Genomes Project, we observed that pathogenic *HTT* CAG expansions were two- to three-fold more frequent than expected from clinical prevalence estimates, suggesting incomplete penetrance or under-diagnosis, particularly among individuals with shorter pathogenic alleles^7^.

Advanced neuroimaging can detect structural and functional brain changes years before clinical diagnosis of HD^8^, with pre-symptomatic volumetric changes predicting disease course^9^ and informing early disease staging^10^. Population cohorts with sr-WGS, neuroimaging and clinical data now provide an opportunity to interrogate repeat length as a quantitative biological modifier across both clinical and subclinical phenotypes and determine the penetrance of the pathogenic expansions. A recent population-scale survey across 37 repeat-expansion loci confirmed that pathogenic *HTT* carriers show disease-associated striatal volume loss prior to diagnosis^11^, but did not examine continuous repeat-length effects within the normal and intermediate range.

Here, we investigate *HTT* CAG repeat length as a continuous biological variable in UK Biobank, a large population-based cohort of ∼500,000 individuals recruited outside clinical settings, providing an unselected sample of the general population. We test whether variation within the normal and intermediate range is associated with differences in brain structure, cognition, and neuropsychiatric risk, and quantify age-specific penetrance among carriers of pathogenic repeat expansions.

## 3. Methods

### 3.1 UK Biobank Cohort

Participants were drawn from UK Biobank, a large prospective cohort study which recruited over 500,000 adults aged 40–69 years across the United Kingdom between 2006 and 2010. At baseline assessment, participants provided detailed health, sociodemographic and lifestyle information and completed touchscreen-based cognitive assessments. Neuroimaging data were collected for a subset of participants at two further timepoints.

All participants provided informed consent through electronic signature at baseline assessment. The UK Biobank project was approved by the Northwest Haydock Research Ethics Committee (reference: 11/NW/0382).

### 3.2 Short Tandem Repeat Genotyping

The CAG repeat lengths in the *HTT* gene were obtained for 474,446 individuals from the UK Biobank using ExpansionHunter version 5.0.0^12^. Variant call files were processed in R within the UK Biobank Research Analysis Platform (RAP). For alleles ≥ 36 repeats, repeat sizes were manually reviewed using pileup plots; calls were accepted if supported by one or more spanning reads or 3 or more flanking reads consistent with the expansion. The longer allele (A2) was used as the primary measure of repeat length throughout the analyses. The custom code for the applets used to generate and annotate the *HTT* repeat sizes is available at: https://github.com/chrisclarkson/ukbb_str_pipeline.

### 3.3 Imaging Analysis

#### 3.3.1 Neuroimaging Measures

Neuroimaging was performed at three UK Biobank imaging centres (Newcastle upon Tyne, Stockport, and Reading) using identical Siemens Skyra 3T scanners (software version VD13) equipped with a standard 32-channel head coil. Analyses used image-derived phenotypes derived from T1-weighted structural MRI acquired at the first UK Biobank imaging visit.

Full details of the UK Biobank imaging pre-processing and quality control pipeline have been described previously^13^. Briefly, T1-weighted images were corrected for gradient distortions and registered to Montreal Neurological Institute (MNI) space using FLIRT and FNIRT. Brain extraction was performed using BET, followed by tissue-type segmentation using FAST.

Subcortical volumes were obtained from UK Biobank imaging-derived phenotypes generated using FSL FIRST, while global tissue and compartment volumes (grey matter, white matter, and CSF/ventricular measures) were derived from the standard UK Biobank T1 structural imaging pipeline.

#### 3.3.2 Imaging Cohort

59,717 individuals who passed *HTT* CAG genotyping QC had structural MRI data acquired at the first UK Biobank imaging visit (undertaken between 2014 and 2023). Analyses were restricted to participants of White British ancestry, as defined by UK Biobank using concordant self-reported ethnicity and genotype-based principal component clustering, retaining 50,677 individuals. Related individuals up to third degree were identified using the UK Biobank kinship file, and one individual from each related pair was randomly excluded, yielding an unrelated subset of 48,939 individuals (1,738 removed).

Imaging and covariate quality control were performed in two stages. First, individuals with extreme covariate values, including head motion and volumetric scaling (>5 SD from the sample mean), were excluded (n = 184), leaving 48,755 individuals. Individuals with extreme global brain-volume measures (>5 SD) were then excluded (n = 142), resulting in a sample of 48,613 participants. Extreme subcortical volume values (>5 SD) were set to missing rather than excluded at the individual level.

European ancestry–specific principal components were derived from genotype data. Individuals without available or high-quality genotype data were excluded (n = 235), resulting in a final sample of 48,378 participants. Participants with missing covariates were excluded on a per-analysis basis.

Within the final dataset, 45,150 individuals carried normal *HTT* alleles, 3,115 carried intermediate alleles, 107 carried reduced-penetrance alleles, and 6 carried pathogenic alleles (see **Supplementary Figure SF1** for cohort flow diagram). All analyses were conducted in R (version 4.2.2).

#### 3.3.3 Statistical analysis of repeat-length effects and summary metrics

Based on prior evidence that HD consistently affects subcortical and global brain volumes^10,9^, with the strongest and most consistent effects observed in the caudate, pallidum, putamen, thalamus, ventricular volume, and total white matter volume^10^, we examined seven bilateral subcortical volumes (amygdala, accumbens, caudate, pallidum, putamen, thalamus, and hippocampus) and seven global volumetric measures (ventricular CSF, total CSF, subcortical grey matter, cortical grey matter, total grey matter, total white matter, and total brain volume) derived from structural MRI at the first UK Biobank imaging visit. For each IDP, the predictor of interest was the *HTT* CAG repeat length of the longer *HTT* allele.

Linear regression models were fitted separately for each brain volume to estimate the association between *HTT* CAG repeat length and brain volume within the normal and intermediate repeat range (CAG ≤35). Models were adjusted for head-size scaling (volumetric scaling factor), head motion (mean resting-state fMRI head motion, log-transformed, and T1 structural motion), scanner position (brain position X/Y/Z and table position), imaging centre, time between baseline assessment and imaging, age (modelled using a second-degree orthogonal polynomial), sex, an age-by-sex interaction term, and genetic ancestry (top ten European-specific principal components). All continuous predictors were standardised prior to modelling, and residuals were approximately normally distributed on visual inspection. All statistical analyses were conducted in R (version 4.2.2).

To characterise repeat-length–associated effects beyond the primary linear model, we defined two complementary metrics: age-dependent modulation of CAG effects within the normal and intermediate range, and deviation at longer repeat lengths (CAG ≥ 36).

Age-dependent modulation was assessed for CAG ≤35 using an interaction between CAG repeat length and the first component of the age polynomial. This was complemented by visualisation of age-stratified linear fits of residualised volume on CAG within the same range, with individuals split at the median age (≤66 vs ≥67 years).

Deviation at reduced-penetrance and pathogenic repeat lengths (CAG ≥36) was assessed by testing departure from the linear relationship defined in the normal and intermediate range (CAG ≤35). Volumes were first residualised using a covariate-only model (excluding repeat length), after which a linear model of residualised volume versus CAG was fitted for CAG ≤35 and extrapolated to larger repeat lengths. Deviation was defined as the mean difference between observed and predicted residuals in individuals with CAG ≥36 and tested against zero using a one-sample t-test. This approach identifies non-linearity relative to the empirically observed linear trend in the ≤35 range. All p values were corrected for multiple comparisons using the Benjamini–Hochberg false discovery rate (FDR) procedure across all imaging phenotypes and summary metrics.

For visualisation, CAG repeat lengths were grouped into predefined bins across the full allelic range and mean residualised volume (±SE) was plotted against mean repeat length per bin. The linear trend estimated within CAG ≤35 was overlaid and extrapolated beyond 35 repeats to provide a visual reference for reduced-penetrance and pathogenic alleles.

### 3.4 Clinical Phenotypes: Depression, Anxiety, Delirium and Dementia

To explore the population-level effects of *HTT* CAG repeat length across domains of brain and behavioural function, we analysed psychiatric, clinical, and cognitive phenotypes in UK Biobank. Analyses focused on individuals with normal and intermediate repeat lengths to define baseline population trends, with comparison to reduced penetrance and pathogenic alleles to identify deviations from these relationships. Associations were examined using both continuous repeat length and predefined repeat-length categories: normal (CAG ≤26), intermediate (CAG 27–35), and reduced-penetrance/pathogenic (CAG ≥36), with phenotype-specific analytical approaches as detailed below.

#### 3.4.1 Depression

The analysis cohort comprised individuals with available *HTT* CAG repeat-length data who had completed at least one UK Biobank Mental Health Questionnaire (MHQ1 or MHQ2). Depression status was coded as present if participants endorsed “Depression” in response to the MHQ item “Have you been diagnosed with one or more of the following mental health problems by a professional, even if you don’t have it currently (tick all that apply)?” in either MHQ1 (Field 20544) or MHQ2 (Field 29000), capturing lifetime clinically recognised depression. Participants who completed one or both questionnaires and did not report depression were classified as unaffected and right censored at the age of their most recent MHQ completion. Among individuals with *HTT* repeat-length data, 149,356 completed MHQ1 and 166,290 completed MHQ2; 207,428 completed at least one questionnaire and 108,218 completed both.

Depression status was defined using self-reported professional diagnosis from the two UK Biobank Mental Health Questionnaires (MHQ). This approach captured a lifetime depression prevalence of approximately 21%, closer to general population lifetime prevalence estimates than linked healthcare records, which identified depression in only approximately 13% of the cohort, likely reflecting the well-documented under-ascertainment of depression in routine record-based data^14^.

Among MHQ-defined cases, age at onset was defined as the earliest available age from: (i) MHQ1-reported age at first depressive episode, (ii) MHQ2-reported age at first depressive episode, or (iii) ICD-10–derived age based on the first recorded diagnosis of depressive episode (F32) or recurrent depressive disorder (F33). ICD-10 records were used solely to derive onset age. Onset age was derived from MHQ1 in 30,614 cases, MHQ2 in 10,601 cases, and ICD-10 records in 2,216 cases.

After excluding cases without an available onset age (n = 3,425) and individuals with incomplete genetic ancestry data (n = 2,016), the final analytic cohort comprised 201,987 individuals (43,000 affected and 158,987 unaffected).

Time-to-event analyses were conducted using age as the time scale. For affected individuals, survival time was defined as age at first depressive episode; unaffected individuals were right censored at age at most recent MHQ completion. Participants were grouped according to *HTT* CAG-repeat length category, defined as normal (≤26), intermediate (27–35), and reduced-penetrance/pathogenic (≥36).

Kaplan–Meier curves were constructed to visualise age-dependent, depression-free survival across *HTT* CAG-repeat length categories, with differences assessed using log-rank tests. Cox proportional hazards models were fitted with *HTT* CAG-repeat category as the primary exposure, using the normal group as the reference. Models were adjusted for genetic ancestry using the first 10 ancestry principal components derived from genome-wide genotype data. Proportional hazards assumptions were assessed using Schoenfeld residuals; because sex violated this assumption, primary Cox models were stratified by sex. Hazard ratios and 95% confidence intervals were reported. As a complementary analysis, CAG repeat length was also modelled as a continuous variable within the normal and intermediate range (CAG ≤35) to assess graded effects across this range.

#### 3.4.2 ICD-10 outcomes: Dementia, Anxiety and Delirium

Time-to-event analyses were performed for dementia, anxiety, and delirium using linked UK Biobank health record data. Case status and age at onset were derived from ICD-10 first-reported year fields, which aggregate diagnoses across multiple sources including hospital episode statistics, primary care records, death registry data, and self-reported information. Dementia was defined as the earliest recorded diagnosis of Alzheimer’s disease (F00), vascular dementia (F01), dementia in other diseases classified elsewhere (F02), or unspecified dementia (F03). Anxiety was defined by a diagnosis of other anxiety disorders (F41), and delirium by delirium not induced by alcohol or other psychoactive substances (F05). Age at diagnosis was estimated as the difference between the year of first recorded diagnosis and year of birth, and implausible ages were excluded prior to analysis (dementia: <20 years; anxiety: <3 years). Age was used as the time scale; event time for affected individuals was defined as age at first diagnosis, and those without a diagnosis were right-censored at the earlier of age at death or administrative end of follow-up (2025). Participants were grouped by *HTT* CAG repeat length as normal (≤26), intermediate (27–35), and reduced-penetrance/pathogenic (≥36).

Kaplan–Meier curves were used to estimate age-dependent, diagnosis-free survival across *HTT* CAG categories, with differences assessed using log-rank tests. Cox proportional hazards models were fitted with *HTT* CAG category as the primary exposure (normal group as reference), adjusting for genetic ancestry (first 10 genotype-derived principal components). Proportional hazards assumptions were assessed using Schoenfeld residuals; as sex showed evidence of time-varying effects, models were stratified by sex. Hazard ratios (HRs) and 95% confidence intervals were reported. As a complementary analysis, CAG repeat length was also modelled as a continuous variable within the normal and intermediate range (CAG ≤35) using Cox proportional hazards models.

For delirium, an age-restricted landmark analysis was performed to examine risk at older ages. Analyses were restricted to individuals with follow-up beyond age 80 who were delirium-free at that age. Follow-up was re-originated at age 80, and post-80 delirium incidence was compared between the normal and intermediate *HTT* repeat groups using Kaplan–Meier curves and an unadjusted log-rank test.

### 3.5 Cognitive Phenotypes

We selected three UK Biobank cognitive phenotypes, symbol digit substitution, reaction time, and fluid intelligence, to sample complementary domains relevant to adult cognitive function while prioritising measures with reasonable psychometric support and broad availability. These tasks capture key domains, processing speed and higher-order reasoning, that are known to decline with ageing and are particularly relevant to early neurodegenerative change. Reaction time and symbol digit substitution provide sensitive indices of processing speed, while fluid intelligence reflects broader reasoning ability and contributes to the general cognitive factor (“g”). Within UK Biobank, these measures show moderate-to-good reliability^15^, with both symbol digit substitution and fluid intelligence contributing strongly to a general cognitive factor^16^ that captures shared variance across tasks.

Cognitive phenotypes, including reaction time, fluid intelligence, and symbol digit substitution, were analysed using linear regression, with reaction time and symbol digit substitution modelled as continuous outcomes and fluid intelligence, measured on a bounded discrete scale, treated as approximately continuous. For participants with data available at multiple timepoints, the result from the latest available assessment was selected (the assessment at the oldest age). Primary models included *HTT* CAG repeat length of the longer allele as a continuous predictor and were fitted in individuals with repeat lengths in the normal and intermediate range (CAG ≤35), adjusting for age at assessment, modelled as a second-degree orthogonal polynomial, sex, an interaction between sex and the first age polynomial component, and the first ten ancestry principal components. Analyses were restricted to individuals with non-missing CAG repeat length, available cognitive data, and complete covariate information.

To assess whether results at the longest repeat lengths departed from the linear relationship observed within the normal and intermediate range (CAG ≤35), a deviation metric was calculated for individuals with CAG ≥36. Phenotypes were first residualised using a covariate-only model, after which a linear model of residualised phenotype versus CAG was fitted for individuals with CAG ≤35 and extrapolated to larger repeat lengths. Deviation was defined as the mean difference between observed and predicted residuals in individuals with CAG ≥36 and tested against zero using a one-sample t-test. This approach identifies non-linearity relative to the empirically observed linear trend in the ≤35 range. For visualisation, mean residualised phenotype values (±SE) were calculated within CAG repeat-length bins across the full allelic range.

### 3.6 Population-based penetrance and Huntington’s Disease ascertainment

#### 3.6.1 Ascertainment of HD diagnosis and age dependent penetrance

Participants with *HTT* CAG repeat length ≥36 in the longer allele (A2) were identified. Carrier frequencies for 36–39 (reduced-penetrance) and ≥40 (pathogenic) repeats were calculated relative to all genotyped individuals with 95% confidence intervals estimated using binomial tests. For age-dependent diagnosis analyses, repeat length was grouped into 36–37, 38*–*39, 40–41, and ≥42 repeats. HD diagnosis was defined by the ICD-10 code G10 derived from linked health records aggregating diagnoses across hospital episode statistics, primary care records, and death registry data. Age at diagnosis was calculated as the year of first recorded G10 code minus year of birth. Kaplan–Meier analyses used age as the time scale, with the event defined as first recorded G10 diagnosis; individuals without a G10 code were right-censored at last available follow-up (administrative censoring in 2025). Kaplan–Meier curves were compared descriptively across repeat-length categories.

#### 3.6.2 Imaging Staging for Pathogenic Allele Carriers

To contextualise disease burden across individuals with different CAG repeat lengths and ages, we used the CAG-Age Product (CAP) score, a widely used composite metric that integrates age and repeat length to approximate cumulative exposure to mutant huntingtin^17^ providing a biologically informed index of disease progression that facilitates comparison across cohorts with differing ascertainment strategies. We used a scaled CAP metric (CAP100), where values approaching 100 have been associated with transition toward manifest disease^18^, enabling comparison with established clinical staging frameworks.

Participants carrying reduced penetrance and pathogenic *HTT* CAG repeat lengths (CAG ≥36) that had accompanying MRI neuroimaging were assessed for possible Stage 1 disease using the Huntington’s Disease Integrated Staging System (HD-ISS), proposed by Tabrizi et al^10^. Caudate and putamen volumes were normalised to intracranial volume and compared with age-specific thresholds; individuals with adjusted volumes below threshold were classified as meeting imaging criteria for Stage 1 disease. To characterise the relationship between genetic burden and imaging abnormality, associations between CAP100 and continuous striatal deviation from age-specific thresholds were evaluated using linear regression models.

To compare disease burden at imaging-defined Stage 1 between UK Biobank and clinically ascertained cohorts, CAP100 values were examined in UK Biobank participants meeting Stage 1 criteria and stratified by repeat-length category (reduced penetrance, CAG 36–39; pathogenic, CAG 40–42). These distributions were compared with corresponding Stage 1 CAP100 distributions from clinical cohorts, approximated by simulation from published summary statistics (mean, standard deviation, and sample size) reported in Long *et al*. ^19^. Long *et al*.^19^ applied the HD-ISS to a large multi-study HD dataset (Enroll-HD, IMAGE-HD, PREDICT-HD, TRACK-HD/ON) using a machine learning imputation algorithm to assign stages where imaging landmark variables were unavailable. Summary statistics were combined across CAG repeat lengths within each allele group using weighted means and pooled standard deviations prior to simulation.

#### 3.6.3 ICD-10 phenotypic enrichment in pathogenic *HTT* allele carriers

ICD-10 phenotypes were identified from first-reported year fields, which aggregate diagnoses across multiple sources including hospital episode statistics, primary care records, death registry data, and self-reported information. Phenotypes were binarized as ever/never. Analyses were restricted to individuals with *HTT* CAG repeat lengths ≥40 (pathogenic) or ≤26 (normal), excluding Huntington’s disease diagnoses phenotype (ICD-10 G10).

We performed a primary analysis including all pathogenic carriers and a sensitivity analysis excluding those with a recorded Huntington’s disease diagnosis. Phenotypes were required to have ≥20 cases overall and ≥5 cases in both groups; in the sensitivity analysis, ≥3 cases in pathogenic carriers were permitted due to reduced sample size. Logistic regression models were fitted with pathogenic repeat status as the exposure, adjusting for age, age², sex, age×sex interaction, and ten ancestry principal components; odds ratios (ORs) and 95% confidence intervals were derived from model coefficients. Multiple testing was controlled using the Benjamini–Hochberg false discovery rate.

#### 3.6.4 Cognitive Phenotypes for Pathogenic Allele Carriers

Cognitive performance was examined using reaction time, fluid intelligence, and symbol digit substitution measures derived from UK Biobank assessments. For participants with data available at multiple assessment instances, the latest available assessment was selected. To quantify deviation from expected performance, a normative modelling framework was applied using a reference group restricted to individuals with normal *HTT* repeat length (CAG ≤26).

Within this reference group, covariate-only linear regression models were fitted separately for each cognitive phenotype, adjusting for age at assessment (modelled as a second-degree orthogonal polynomial), sex, an interaction between sex and the first age polynomial component, and the first ten ancestry principal components. Predicted values were generated for all participants, and residuals were standardised using the residual standard deviation from the reference model to yield normative z-scores.

Normative deviation was summarised by estimating the proportion of individuals exceeding predefined z-score thresholds (1, 1.645, and 2 standard deviations from the reference mean), with the 1.645 SD threshold corresponding to the upper 5% of the reference distribution. These thresholds were used to characterise the extent to which pathogenic *HTT* carriers fell within or outside the expected range of cognitive performance across age and repeat-length groups.

## 4. Results

### 4.1 *HTT* CAG repeat length below the pathogenic range is associated with decreased brain volumes

To determine whether repeat-length variation within the normal and intermediate range is associated with measurable differences in brain volume, we analysed 48,265 individuals from UK Biobank with *HTT* CAG repeat lengths ≤35 (45,150 with normal alleles and 3,115 with intermediate alleles) using linear regression models adjusted for head size, motion, position in scanner, age, sex and genetic ancestry.

We observed significant negative associations between *HTT* CAG repeat length and total volume of the nucleus accumbens (β = −0.0127, pFDR = 7.4 × 10⁻³), putamen (β = −0.0110, pFDR = 9.3 × 10⁻³), thalamus (β = −0.0091, pFDR = 1.6 × 10⁻²), and hippocampus (β = −0.0120, pFDR = 1.1 × 10⁻²) (**Figure 2 and Supplementary Figures SF2 and SF3)**. No significant associations were seen with the amygdala, caudate or globus pallidus.

**Figure 1:**
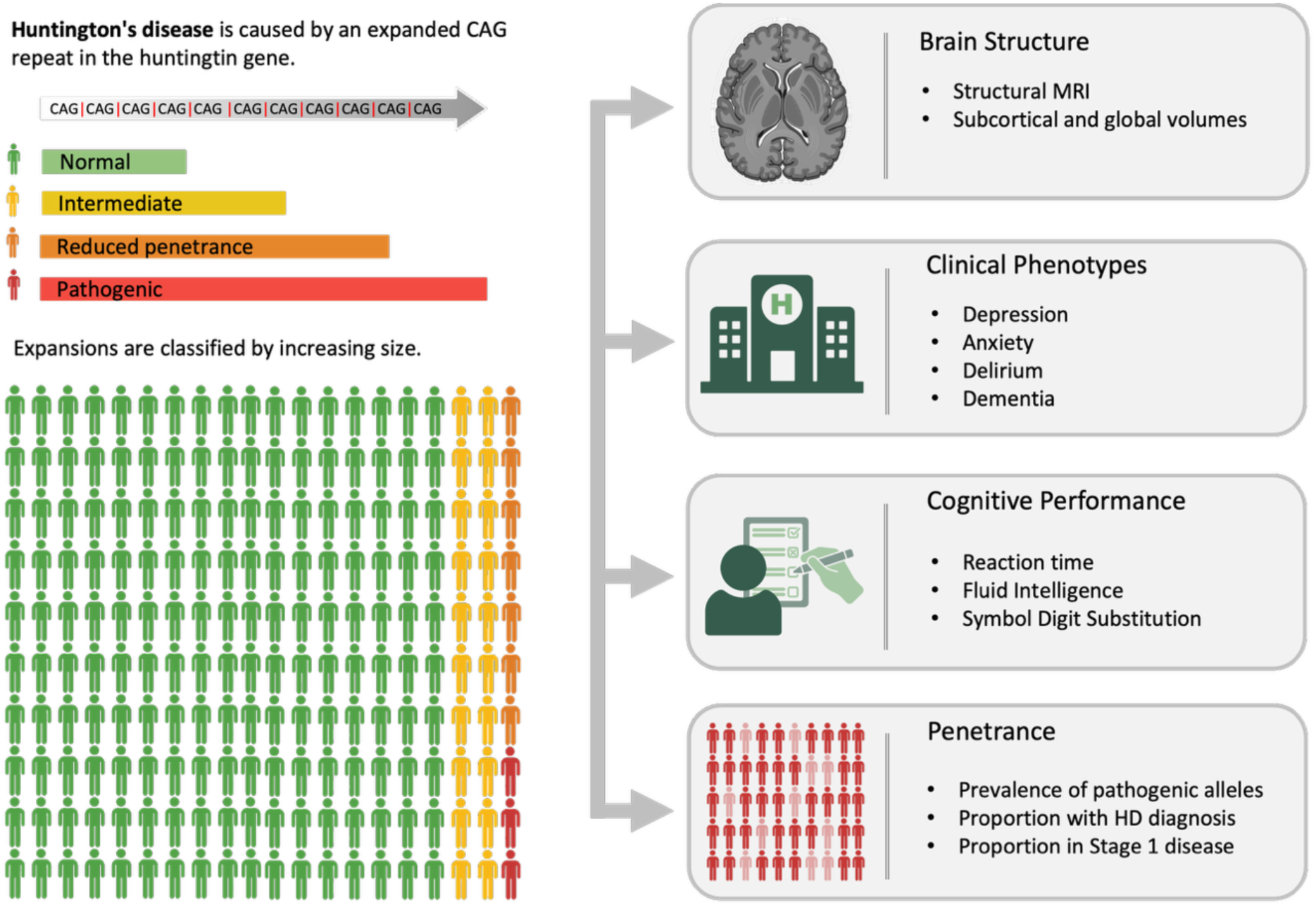
Study Overview: Huntingtin CAG repeat length is tested as a continuous modifier of brain structure and health vulnerability in a large human population.

**Figure 2.**
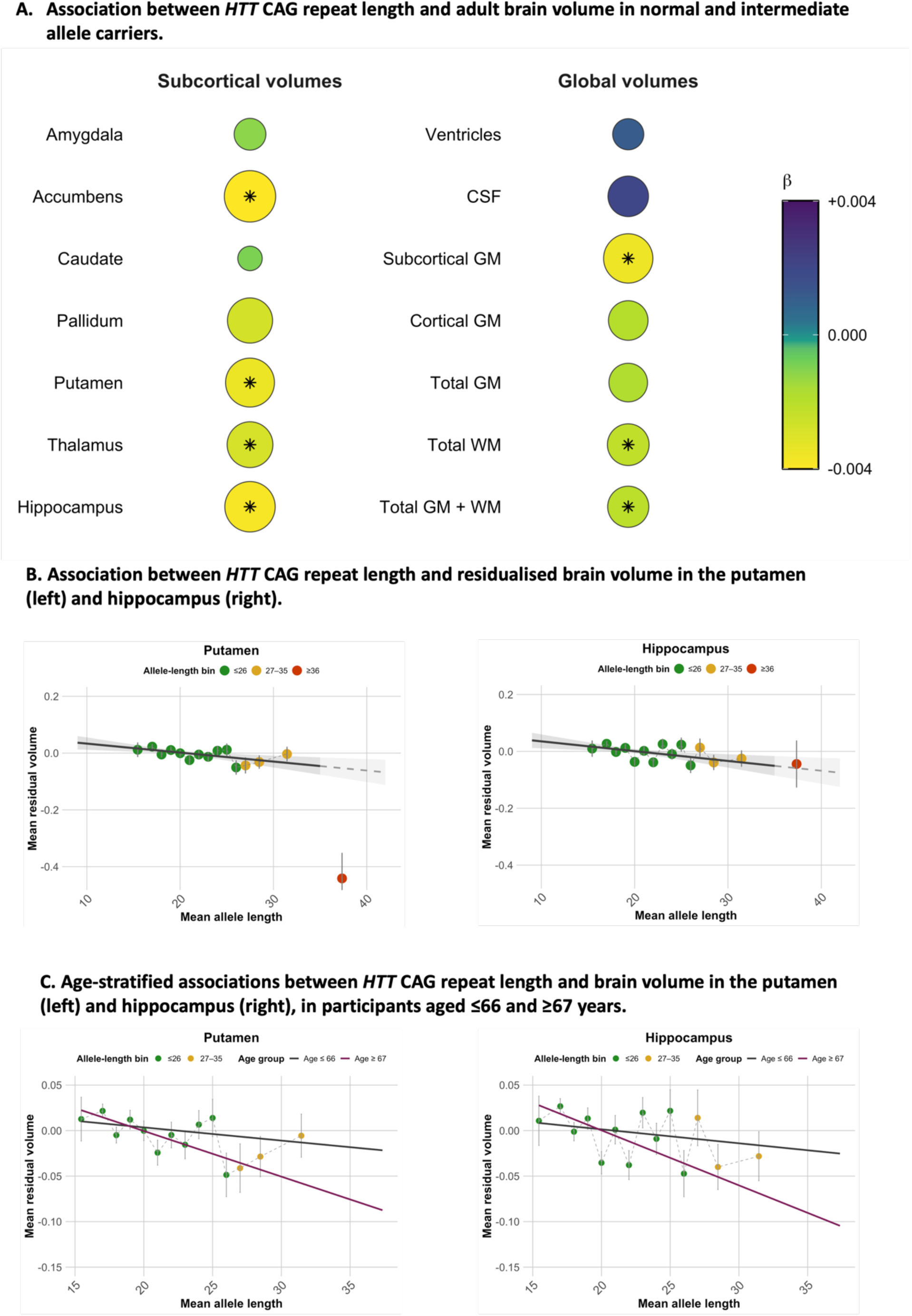
Association between HTT CAG repeat length (longer allele) and brain volumes in UK Biobank. **(A)** Rows show standardised subcortical (left) and global (right) volumes; colour indicates effect size, circle size reflects magnitude, and outlines denote statistical significance. **(B)** Association between HTT CAG repeat length and residualised subcortical volume in the putamen (left) and hippocampus (right). Points represent mean residuals within CAG bins (±SE), coloured by allele range (normal ≤26, intermediate 27–35, reduced-penetrance/pathogenic ≥36). The regression line (±95% CI) is fitted within the normal and intermediate range (≤35) and extrapolated into the pathogenic range. **(C)** Age-stratified equivalent of B. Points represent mean residuals within CAG bins (±SE), coloured by allele range (normal ≤26, intermediate 27–35). Linear fits shown separately for participants aged ≤66 years (dark grey) and ≥67 years (maroon), restricted to CAG ≤35.

At the global level, longer *HTT* CAG repeat length was also associated with reduced total subcortical grey matter volume (β = −0.0113, pFDR = 1.7 × 10⁻³), cortical grey matter volume (β = −0.0061, pFDR = 2.8 × 10⁻²), total grey matter volume (β = −0.0057, pFDR = 2.6 × 10⁻²), total white matter volume (β = −0.0070, pFDR = 9.3 × 10⁻³), and combined grey and white matter volume (β = −0.0067, pFDR = 3.4 × 10⁻³) (**Supplementary Figures SF4 and SF5**). No significant association was observed for ventricular or total cerebrospinal fluid volumes.

Effect sizes were modest but consistent in direction across affected structures, indicating a distributed association between increasing *HTT* CAG repeat length and reduced brain volume in the normal and intermediate range.

To formally assess age-dependent modulation, age × CAG interaction terms were included in fully adjusted models within the normal and intermediate range with a significant interaction observed for the nucleus accumbens. Age-stratified analyses supported a broader pattern of age-related amplification across all 14 imaging phenotypes, with larger effect in older participants (≥67 years) than in younger individuals in every case (exact binomial test p = 1.2 × 10⁻⁴, **Figure 2C and Supplementary Figure SF6 and SF7**), suggesting a systematic strengthening of CAG-associated volumetric differences in later life.

Among individuals with reduced-penetrance or pathogenic repeat lengths (≥36 repeats), significant deviations from the normal/intermediate linear trend were observed for the putamen, caudate, and nucleus accumbens, but not for the amygdala, thalamus, or hippocampus, where volumes for the longest repeat lengths aligned closely with the extrapolated linear trend (**Figure 2B, Supplementary Figures SF2 and SF3**). The pallidum showed nominal evidence of deviation that did not survive multiple-testing correction. For global phenotypes, significant deviations were observed across nearly all measures, with total CSF volume being the only exception; significant effects were seen for total brain volume, cortical and total grey matter, total white matter, subcortical grey matter, and ventricular volume (**Supplementary Figures SF4 and SF5**). The deviations observed in individuals with reduced-penetrance or pathogenic alleles were most pronounced in striatal structures, consistent with their recognised susceptibility to HTT-mediated neurodegeneration.

### 4.2 Clinical Phenotype Results: Depression, Anxiety, Delirium and Dementia

Using Mental Health Questionnaire data (201,987 participants; 43,000 individuals with depression), we observed a graded association between *HTT* CAG repeat length and depression risk (**Figure 3A**). Compared with the normal range, individuals with intermediate repeats had a modestly increased risk (HR = 1.054, 95% CI 1.015–1.096, pFDR = 0.026). Those with reduced-penetrance/pathogenic repeats showed a larger effect size (HR = 1.220, 95% CI 1.012–1.472), although this did not survive multiple testing correction (pFDR = 0.074).

**Figure 3.**
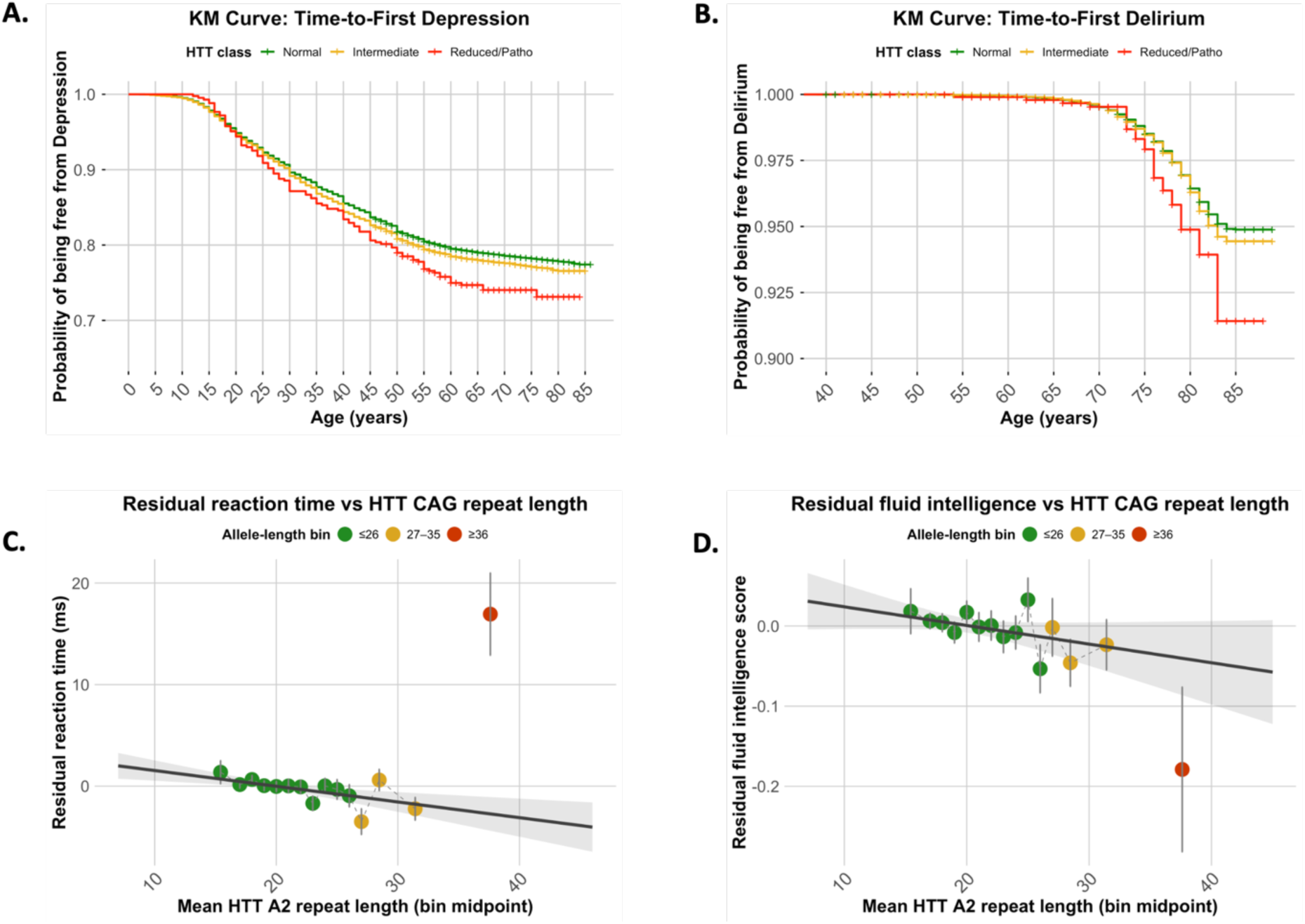
Top row: Kaplan–Meier curves showing age-dependent depression-free survival (A) and delirium-free survival (B) across three HTT CAG repeat length categories: normal (≤26, green), intermediate (27–35, yellow), and reduced-penetrance/pathogenic (≥36, red). Bottom row: associations between HTT CAG repeat length and residualised cognitive performance for Reaction Time (C), Fluid Intelligence (D), and Symbol Digit Substitution (E). Points represent mean residuals within CAG bins (±SE), coloured by allele range (normal ≤26, intermediate 27–35, reduced-penetrance/pathogenic ≥36). The regression line (±95% CI) is fitted within the normal and intermediate range (≤35) and extrapolated into the pathogenic range.

Across ICD-10–defined outcomes (∼469,000 participants; 39,788, 8,529 and 8,645 individuals with anxiety, delirium and dementia respectively, **Supplementary Table ST2**), associations differed by phenotype. There was no evidence of association with anxiety (F41) (**Supplementary Figure SF8 B)**, with hazard ratios close to unity for both intermediate (HR = 1.01, 95% CI 0.97–1.05, pFDR = 0.74) and reduced-penetrance/pathogenic repeats (HR = 1.08, 95% CI 0.89–1.33, pFDR = 0.49). Delirium (F05) showed increased risk among individuals with reduced-penetrance/pathogenic repeats (HR = 1.55, 95% CI 1.08–2.21, pFDR = 0.045), but not for intermediate repeats (**Figure 3B**). However, Kaplan–Meier curves suggested divergence at older ages, with borderline evidence of increased post-80 risk in intermediate carriers (log-rank p = 0.05). Dementia showed the strongest association restricted to reduced-penetrance/pathogenic carriers (HR = 1.81, 95% CI 1.30–2.52, pFDR = 0.0037). No association was observed for intermediate repeats (**Supplementary Figure SF8 A**). Within the normal and intermediate range, continuous modelling showed modest associations with depression (HR per repeat 1.003, 95% CI 1.000–1.006, p = 0.045) and delirium (HR 1.007, 95% CI 1.001–1.013, p = 0.022), but not anxiety or dementia. Full results and sample sizes are provided in **Supplementary Tables ST1 and ST2**.

### 4.3 Cognitive Phenotype Results: reaction time, fluid intelligence and symbol digit substitution test

Within the normal and intermediate *HTT* CAG-repeat range (≤35 repeats), repeat length was significantly associated with reaction time (**Figure 3C**), with longer repeats associated with faster performance (β = −0.155, pFDR = 0.0067, n=463,375, mean age = 57.9 years, SDage = 8.5 years). In contrast, fluid intelligence showed a weaker trend in the opposite direction (**Figure 3D**), with lower scores at longer repeat lengths that did not reach statistical significance in the full sample (β = −0.0023, pFDR = 0.096, n= 205,837, mean age = 59.9, SDage = 8.9 years), and symbol digit substitution showed no evidence of association (β = −0.0067, pFDR = 0.096, n = 114,929, mean age = 56.6 years, SDage= 7.8 years).

At longer repeat lengths (≥36 repeats), domain-specific patterns emerged. Reaction time showed a clear deviation from the linear relationship observed in the normal and intermediate range (Δ = 19.7 ms, pFDR = 7.9 × 10⁻⁶), with reversal of the normal-range direction. Neither fluid intelligence nor symbol digit substitution deviated significantly from the extrapolated trend (both pFDR = 0.21) with fluid intelligence remaining directionally consistent with its normal-range association.

Analyses restricted to individuals of European ancestry were consistent, with the reaction time association (β = −0.150, pFDR = 0.018) and deviation at longer repeat lengths (Δ = 21.2 ms, pFDR = 2.3 × 10⁻⁵) both remaining significant, and the fluid intelligence association within the normal range reaching significance (β = −0.0038, pFDR = 0.033). Full results are in **Supplementary Table ST3**.

### 4.4 Population-based penetrance

#### 4.4.1 Huntington’s Disease Diagnosis in Reduced penetrance and Pathogenic Allele Carriers

We identified all UK Biobank participants carrying *HTT* CAG repeat alleles in the reduced penetrance and pathogenic range (≥36 repeats; N = 1028). The carrier frequency for repeat length in the pathogenic range (CAG ≥40) was 1 in 3060 (95% CI: 1 in 2616 to 1 in 3605), and 1 in 543 (95% CI: 1 in 509 to 1 in 581) for repeat lengths in the reduced penetrance range (CAG 36–39).

Consistent with the established inverse relationship between repeat length and age at onset^3^, carriers with ≥42 repeats demonstrated the earliest and highest cumulative diagnosis rate (85% by age 75), whereas individuals with 36–39 repeats showed the lowest proportion diagnosed across the observed age range (**Figure 4A**). Among carriers with 40–41 repeats, cumulative diagnosis reached 42% by age 84 (95% CI 19–59%), well below the >95% penetrance predicted for 40 repeats and near-complete penetrance predicted for 41 repeats by current clinical models^20^.

**Figure 4:**
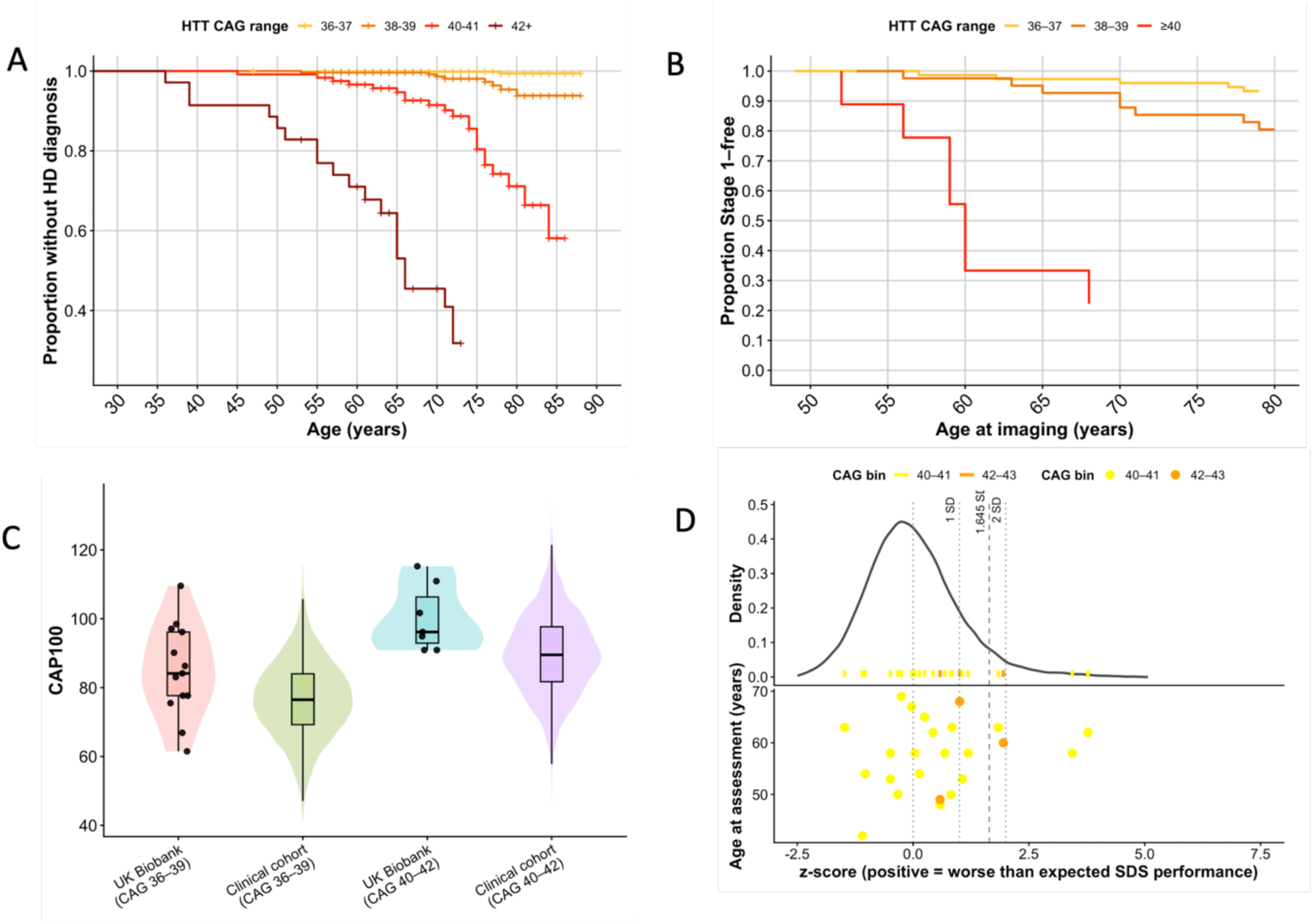
(A) Cumulative proportion of individuals diagnosed with HD (ICD10 G10) by repeat lengths (in 4 categories: 36–37, 38–39, 40–41 and 42+ CAG repeats) (B) Cumulative proportion of individuals remaining free of Stage 1 disease by HTT CAG repeat length (36–37, 38–39 and 40–42 repeats). (C) Comparison of CAP100 distributions of individuals in Stage 1 disease from UK Biobank and clinical cohorts. (D) Normative deviation in Symbol Digit Substitution performance among carriers of pathogenic HTT CAG repeat expansions stratified by CAG bins (40–41, 42–43) for descriptive display.

#### 4.4.2 Imaging Disease Stage for Reduced Penetrance/Pathogenic Allele Carriers

Of the nine pathogenic allele carriers (≥40 repeats) with available structural MRI data, seven (78%) met imaging criteria for at least Stage 1 Huntington’s disease (**Figure 4B**). CAP100 values among UK Biobank participants meeting Stage 1 criteria were consistently higher than those derived from clinically ascertained cohorts (**Figure 4C**). For pathogenic alleles (CAG 40–42) in Stage 1 disease, mean CAP100 was 100.1 (range 90.9–115.3) in UK Biobank compared with 89.7 in clinical cohorts (difference ∼10 units, ∼0.9 SD). For reduced-penetrance alleles (CAG 36–39) in Stage 1 disease, mean CAP100 was 84.9 in UK Biobank compared with 76.6 in clinical cohorts (difference ∼8 units, ∼0.74 SD). As CAP100 incorporates age, these higher values suggest that UK Biobank participants reach Stage 1 imaging criteria at older ages than clinically ascertained individuals; this difference is likely conservative given the tendency of the Long et al.^19^ imputation algorithm to under-assign Stage 1 at lower CAP100 values. None of the UK Biobank Stage 1 participants had a hospital-coded HD diagnosis at the time of analysis.

#### 4.4.3 ICD-10 phenotypic enrichment in pathogenic *HTT* allele carriers

Phenome-wide analysis identified enrichment of neurodegenerative, aspiration-related, and neuropsychiatric diagnoses in pathogenic *HTT* CAG repeat carriers (**Figure 5**). Excluding individuals with a recorded Huntington’s disease diagnosis markedly attenuated these associations, indicating that these are largely driven by those with clinically recorded disease; however, broader neurodegenerative diagnoses remained significant, suggesting that some associations are detectable in individuals without a recorded diagnosis.

**Figure 5.**
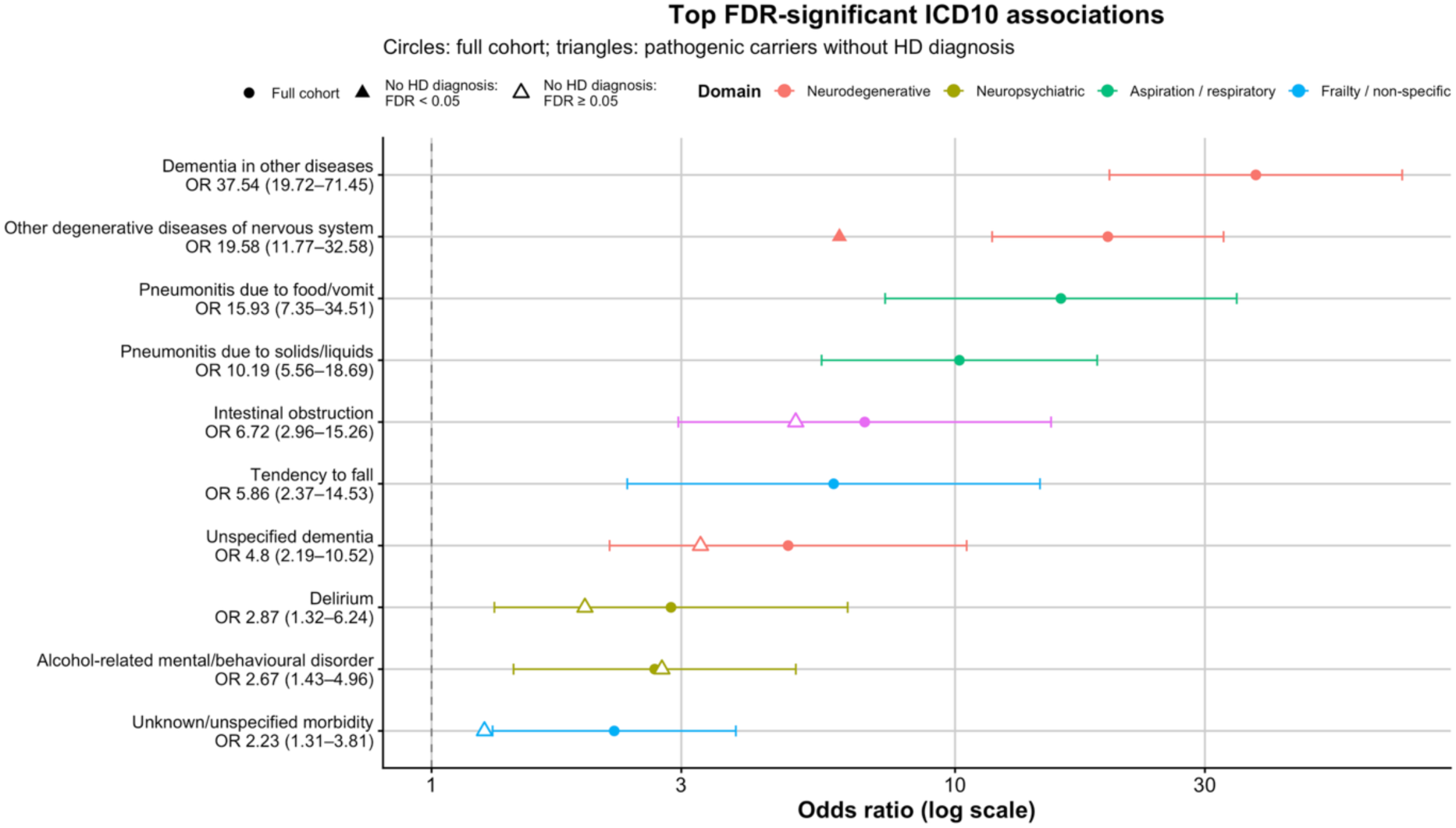
ICD-10 phenotype enrichment associated with pathogenic HTT CAG repeat length in UK Biobank. Forest plot of the FDR–significant ICD-10 phenotypes associated with pathogenic HTT repeat length (CAG ≥40) compared with normal alleles (≤26). Points show odds ratios (ORs) with 95% confidence intervals Circles represent estimates in the full cohort; triangles show estimates after excluding individuals with a recorded Huntington’s disease diagnosis, with filled symbols indicating FDR-significant associations. Phenotypes are grouped by clinical domain.

#### 4.4.4 Cognitive Phenotypes for Pathogenic Allele Carriers

Across all three cognitive measures, the majority of pathogenic allele carriers (CAG ≥40) performed within the expected normative range. For reaction time (n = 149), 82.8% remained below the +1.645 SD threshold, with 17.2% exceeding it; deviation increased with age and repeat length (**Supplementary Figure SF10 A**). For fluid intelligence (n = 60), 91.5% remained above the −1.645 SD threshold, with 8.5% falling below it (**Supplementary Figure SF10 B**). For symbol digit substitution (n = 23), 82.6% remained below the +1.645 SD threshold, with 17.4% exceeding it (**Figure 4D and Supplementary Figure SF10 C**). Across all measures, fewer than 20% of carriers exceeded HD-ISS aligned thresholds, and most remained within the expected range of cognitive performance for their age.

## 5. Discussion

This work provides population-level evidence that *HTT* CAG repeat length acts as a quantitative modifier of brain structure and psychiatric vulnerability across the general population. The finding that repeat-length variation within the normal and intermediate range is associated with measurable differences in brain volume, depression risk, and cognitive processing challenges the assumption that the biological consequences of *HTT* variation are confined to individuals crossing a discrete pathogenic threshold.

This assumption has been difficult to test until now. Prior studies of intermediate allele carriers were limited to small, frequently clinically ascertained cohorts^21^ underpowered to detect modest quantitative effects in population traits. UK Biobank, with its linked neuroimaging, cognitive and clinical outcome data across the full allelic spectrum, provides the scale to resolve these effects directly.

Our findings challenge the categorical model at both ends of the allelic spectrum. Below the pathogenic threshold, repeat-length variation exerts continuous, measurable effects on brain biology, neuropsychiatric risk and cognitive phenotypes. Among pathogenic allele carriers, population-based diagnostic penetrance is substantially lower than in clinically ascertained cohorts: only approximately one third carry a recorded clinical HD diagnosis. Yet the majority of these carriers (78%) who underwent neuroimaging met Stage 1 criteria based on striatal volumes, indicating that early biological disease is present in these individuals. Those reaching imaging-defined Stage 1 appear to do so at an older age than counterparts in clinical cohorts, with CAP100 values approximately 0.7–0.9 standard deviations higher at equivalent repeat lengths. Under-ascertainment within UK Biobank likely contributes to lower observed diagnostic penetrance, but these imaging findings suggest that disease expression in population cohorts may also be genuinely shifted later, particularly among individuals with shorter pathogenic repeat lengths. Together, these observations support a model in which *HTT* CAG repeat length exerts graded, dosage-dependent effects on brain biology across the full allelic spectrum, rather than operating as a simple Mendelian switch.

### *HTT* repeat length shapes brain structure across the population

Recent population-scale studies have demonstrated that pathogenic repeat expansions are associated with brain volume loss prior to clinical diagnosis^11^. Here we extend these observations, showing that variation in *HTT* CAG repeat length is associated with differences in brain structure across the full allelic spectrum.

Across multiple subcortical and global brain structures, longer *HTT* CAG repeat length within the normal and intermediate range was associated with smaller brain volumes. Although modest in magnitude, these effects were highly uniform in direction, supporting a continuous rather than threshold-based model in which repeat-length variation below the pathogenic threshold exerts measurable effects on brain structure. Regional specificity was apparent: putamen and accumbens volumes showed the strongest associations, whilst other structures showed weaker or absent effects, a pattern that broadly mirrors the selective vulnerability of these regions to *HTT*-mediated neurodegeneration at pathogenic lengths. Notably, no association with caudate volume was observed in the normal and intermediate range, despite the caudate’s established vulnerability in manifest HD. This regional dissociation may reflect genuine differences in the sensitivity of these structures to sub-pathogenic repeat-length variation, or alternatively differences in volumetric measurement precision across structures in population-based imaging. A similar pattern was reported in the Huntington’s Disease Young Adult Study, in which putamen but not caudate volume reduction survived multiple testing correction in premanifest carriers approximately 24 years from predicted onset^22^, suggesting that greater sensitivity of the putamen to *HTT*-related effects may extend across the full allelic spectrum.

One plausible explanation for these observations is low-level somatic expansion operating across the lifespan. Handsaker *et al.* ^23^ demonstrated that in pathogenic allele carriers, *HTT* CAG repeats expand somatically in striatal projection neurons over decades, a stochastic process beginning from the inherited germline length and reaching toxic repeat lengths only later in life. Somatic expansion has also been observed in intermediate allele carriers, with low-level gains of +1 to +3 repeats detected in blood DNA, likely an underestimate of expansion in vulnerable neuronal populations given the well-established tissue-specificity of this process^23,24,25^. Under this model, the modest volumetric differences we observe could reflect a very small fraction of neurons in vulnerable regions undergoing somatic expansion sufficient to impair function or viability, an effect only resolvable at population scale.

An alternative explanation is that germline repeat length influences neurodevelopment directly, such that the volumetric differences we observe reflect differences in peak brain volume established early in life rather than progressive attrition. These mechanisms cannot be distinguished in cross-sectional data. However, age-stratified analyses indicated stronger repeat-length effects on brain volume in older individuals (>67 years) than in younger participants (45–66 years), which is more naturally explained by a cumulative process than by a fixed developmental effect. Formal age × repeat-length interaction models provided limited statistical support, with only the nucleus accumbens surviving correction for multiple testing, likely reflecting limited power given modest effect sizes and the restricted age range of the cohort. The descriptive pattern nonetheless mirrors emerging evidence that somatic expansion continues across the lifespan and may progressively amplify the neurobiological consequences of repeat length over time^23^. Longitudinal data spanning a wider age range, ideally from early adulthood, are needed to resolve the relative contributions of neurodevelopmental and cumulative mechanisms.

For individuals with reduced-penetrance or pathogenic alleles, thalamus and hippocampus volumes aligned closely with the extrapolated normal-range trend, reflecting the slower, more linear degeneration characterising these regions in HD^26^. In contrast, striatal structures showed marked deviation, mirroring their known susceptibility to *HTT*-mediated neurodegeneration^27^. Numbers in this subgroup were small and likely depleted of individuals with advanced disease, but the regional pattern of deviation broadly recapitulates the known topography of HD pathology.

### *HTT* repeat length confers neuropsychiatric risk

*HTT* CAG repeat length was associated with neuropsychiatric risk in a domain-specific pattern. Depression risk increased progressively across repeat categories, with both intermediate and reduced-penetrance/pathogenic repeat lengths associated with higher risk relative to the normal range, consistent with prior population-based evidence linking normal-range repeat variation to depression susceptibility^5^. No association was observed with anxiety, supporting domain-specificity rather than a generalised neuropsychiatric effect. For delirium, categorical analyses identified increased risk only among reduced-penetrance/pathogenic carriers, but continuous modelling within the normal and intermediate range demonstrated a significant graded association, suggesting repeat-length variation below the pathogenic threshold contributes to delirium risk independently of frank HD pathology. Dementia showed the strongest effect, restricted to reduced-penetrance/pathogenic carriers with no detectable association in the intermediate range, though whether this reflects a genuinely threshold-like biological effect or limited power to detect modest effects in intermediate carriers remains unclear.

These results indicate that repeat-length variation has measurable consequences for neuropsychiatric risk in the general population, extending beyond its established role in rare monogenic disease and supporting a model in which *HTT* CAG repeat length acts as a graded modifier of neuropsychiatric vulnerability across a continuum of variation.

### Cognitive phenotypes show domain-specific sensitivity

Cognitive phenotypes show domain-specific sensitivity to *HTT* CAG repeat length variation, mirroring evidence from clinical cohorts that different cognitive domains are not uniformly affected by HTT-related pathology^28^.

Fluid intelligence showed a directionally uniform negative association with increasing repeat length, emerging most clearly in European-ancestry restricted analyses. This accords with the expectation that longer repeats, even within the normal and intermediate range, impose a subtle diffuse neurodegenerative burden, detectable on measures of general cognitive ability^16,15^. Symbol digit substitution, assessed in a comparatively smaller sample, showed no significant association in either direction. Given that symbol digit substitution is a sensitive cognitive marker in clinical HD cohorts and features in the HD-ISS staging criteria, the absence of a significant finding may reflect insufficient sample size rather than a true absence of effect at sub-pathogenic repeat lengths.

Reaction time showed an unexpected pattern: faster reaction time with increasing repeat length in the normal and intermediate range, with reversal of this effect among reduced penetrance and pathogenic allele carriers. This aligns with prior evidence that normal-range *HTT* CAG repeat variation may confer cognitive advantage^29,30,31^ and we speculate that it may reflect a form of antagonistic pleiotropy, whereby increasing repeat length confers modest cognitive benefit while simultaneously predisposing to accelerated biological ageing processes that ultimately manifest as neurodegeneration at pathogenic repeat lengths. We note that our data cannot directly test this age-dependent reversal within the normal range, as our sample is predominantly mid-to-late adulthood and the reversal we observe is across allele categories rather than across age strata within the normal range itself.

Taken together, these findings suggest heterogeneity in the relationship between cognitive phenotypes and *HTT* repeat length variation, echoing the variability in cognitive impairment seen across different domains in premanifest or early HD. Our interpretation is constrained by the inherent limitations of population biobank data: there were no clinical cognitive assessments, tests were also completed at different time points, and sample sizes vary markedly across cognitive phenotypes. Longitudinal data spanning a wider age range would allow a more direct test of whether the direction of repeat-length effects on cognition genuinely evolves over the life course.

### Population data reveal reduced penetrance and delayed expression

Our analysis of pathogenic allele carriers provides population-based insight into the clinical expression of longer *HTT* CAG repeats, complementing the continuous effects observed across the normal and intermediate range. Consistent with our previous findings in the 100,000 Genomes Project, pathogenic *HTT* expansions were substantially more frequent in UK Biobank than expected from clinical prevalence estimates of HD^7^. Among individuals with 40–41 CAG repeats, the cumulative proportion with a recorded HD diagnosis was 42% by age 84, lower than the >95% penetrance predicted for 40 repeats and near-complete penetrance predicted for 41 repeats by established clinical models^20^. Among those who underwent neuroimaging, the majority met biomarker-defined Stage 1 criteria, yet CAP100 values were higher than those inferred from clinically ascertained cohorts, suggesting a later age at biomarker transition in this population-based sample. Many carriers with repeat lengths of 40–42 in mid-to-late adulthood demonstrated cognitive performance within normative expectations.

Taken together, these observations suggest greater phenotypic variability in population cohorts than clinical ascertainment would predict, with evidence of both delayed biomarker transition and incomplete clinical recognition. These findings require cautious interpretation, however, as under-ascertainment of clinical diagnosis and healthy volunteer bias^32^ within UK Biobank are both likely to contribute to lower observed penetrance. Separating true biological variability in disease trajectory from these artefactual effects remains difficult in these data.

### Broader implications

Together, these observations support a model in which *HTT* CAG repeat length functions more analogously to a quantitative risk factor, exerting graded, dosage-dependent effects on brain biology across the full allelic spectrum, rather than operating as a simple Mendelian switch. Effect sizes are modest, but they converge in suggesting that the binary framework currently applied to repeat length is an oversimplification that obscures biologically relevant variation. This has relevance as population-scale sequencing increasingly identifies repeat-length variation in unselected individuals for whom current categorical frameworks offer limited guidance.

Repeat-length variation outside the pathogenic range is not biologically neutral, but nor does it confer disease risk of a magnitude that supports a disease-carrier framing. As precision medicine approaches incorporate polygenic and quantitative genetic information, HTT repeat length may nonetheless complement other risk factors, including polygenic scores, age, and imaging biomarkers, as one continuous variable among many that together shape individual biological trajectory, rather than as a categorical determinant of disease.

Replication in ancestrally diverse cohorts and longitudinal studies to establish whether these associations predict clinical outcomes over time remain priorities. Direct measurement of somatic instability across the full allelic spectrum would further help clarify the mechanisms underlying the modest but directionally coherent effects we observe in the normal and intermediate range.

### Strengths and limitations

Strengths of this study include the population-scale design, the integration of neuroimaging, cognition, and clinical outcomes within a single cohort, and the ability to examine repeat-length effects across the full allelic spectrum in an unselected sample. The use of short-read whole-genome sequencing with manual validation of expanded alleles provides confidence in repeat-length assignment at the longer allele lengths, whilst the scale of UK Biobank provides statistical power to detect modest quantitative effects that would be undetectable in conventional cohorts.

Several limitations should be acknowledged. UK Biobank is a volunteer cohort with well-documented healthy participant bias^32^, meaning carriers of pathogenic alleles with earlier or more severe disease expression are probably underrepresented. Imaging analyses were restricted to individuals of White British ancestry, limiting generalisability to other populations. The cross-sectional design precludes causal inference, and the number of individuals with reduced-penetrance or pathogenic repeat lengths who underwent neuroimaging was small and likely depleted of advanced disease. Clinical outcomes were ascertained from ICD-10 coded healthcare records which likely underestimate true event rates. The cognitive measures available, while broadly valid, are brief and were not designed to detect subtle processing changes relevant to early neurodegeneration.

## 6. Conclusions

These results demonstrate that *HTT* CAG repeat length influences brain structure, mental health and cognitive function across mid to late adulthood, with relevance extending beyond rare monogenic disease. These findings support a reframing of *HTT* repeat length as a continuous biological modifier rather than a categorical determinant of disease, with implications for risk prediction and the interpretation of repeat-length variation in population genomics.

## 7. Data Availability

The UK Biobank data are available from https://www.ukbiobank.ac.uk on application.

## 9 Acknowledgements

**This research has been conducted using the UK Biobank Resource under application number 351363.** H.C. was funded by the King’s Health Partners Centre for Translational Medicine. The views expressed are those of the author(s) and not necessarily those of King’s Health Partners. This work was supported by the UK Medical Research Council (award (MR/S006753/1) to A.T. S.T. was supported by research grant funding from the CHDI Foundation, UK Medical Research Council Therapeutic Genomics CoRE (MR/Z504725/1), UK Dementia Research Institute (principally funded by the UK Medical Research Council), and the Wellcome Trust (223082/Z/21/Z). J.L. would like to acknowledge support from the Cure Huntington’s Disease Initiative.

## 10 Ethics Declaration

This research was conducted using the UK Biobank Resource (application number 351363). UK Biobank received ethical approval from the Northwest Multicentre Research Ethics Committee (21/NW/0157). All participants provided informed consent at the time of enrolment.

