## Supplementary Information for "Huntingtin CAG repeat is a continuous modifier of brain structure and health vulnerability"

This file includes:

**Supplementary Figures:**

1) Supplementary Figure SF1: Flow Chart of Study Population for Imaging.
2) Supplementary Figure SF2: *HTT* CAG repeat length versus residualised subcortical volumes.
3) Supplementary Figure SF3: *HTT* CAG repeat length versus residualised subcortical volumes.
4) Supplementary Figure SF4: *HTT* CAG repeat length versus residualised global brain volumes.
5) Supplementary Figure SF5: *HTT* CAG repeat length versus residualised global brain volumes.
6) Supplementary Figure SF6: Age-stratified association between *HTT* CAG repeat length and residualised subcortical volumes.
7) Supplementary Figure SF7: Age-stratified association between *HTT* CAG repeat length and residualised global volumes.
8) Supplementary Figure SF8: Kaplan–Meier curves showing age-dependent dementia-free survival and anxiety-free survival.
9) Supplementary Figure SF9: Residualised cognitive phenotypes vs *HTT* CAG repeat length in UK Biobank (reaction time, fluid intelligence and symbol digit substitution).

10) Supplementary Figure SF10: Normative deviation in reaction time (A), fluid intelligence (B), and Symbol Digit Substitution (C) among carriers of pathogenic *HTT* CAG repeat expansions.

**Supplementary Tables:**

1) Supplementary Table ST1: Effect sizes for psychiatric phenotypes in full mixed ancestry and European cohorts
2) Supplementary Table ST2: Sample sizes and case counts for psychiatric and clinical outcomes
3) Supplementary Table ST3: *HTT* CAG Repeat Length and Cognitive Measures

**Supplementary Figure SF1:** Flow Chart of Study Population for Imaging

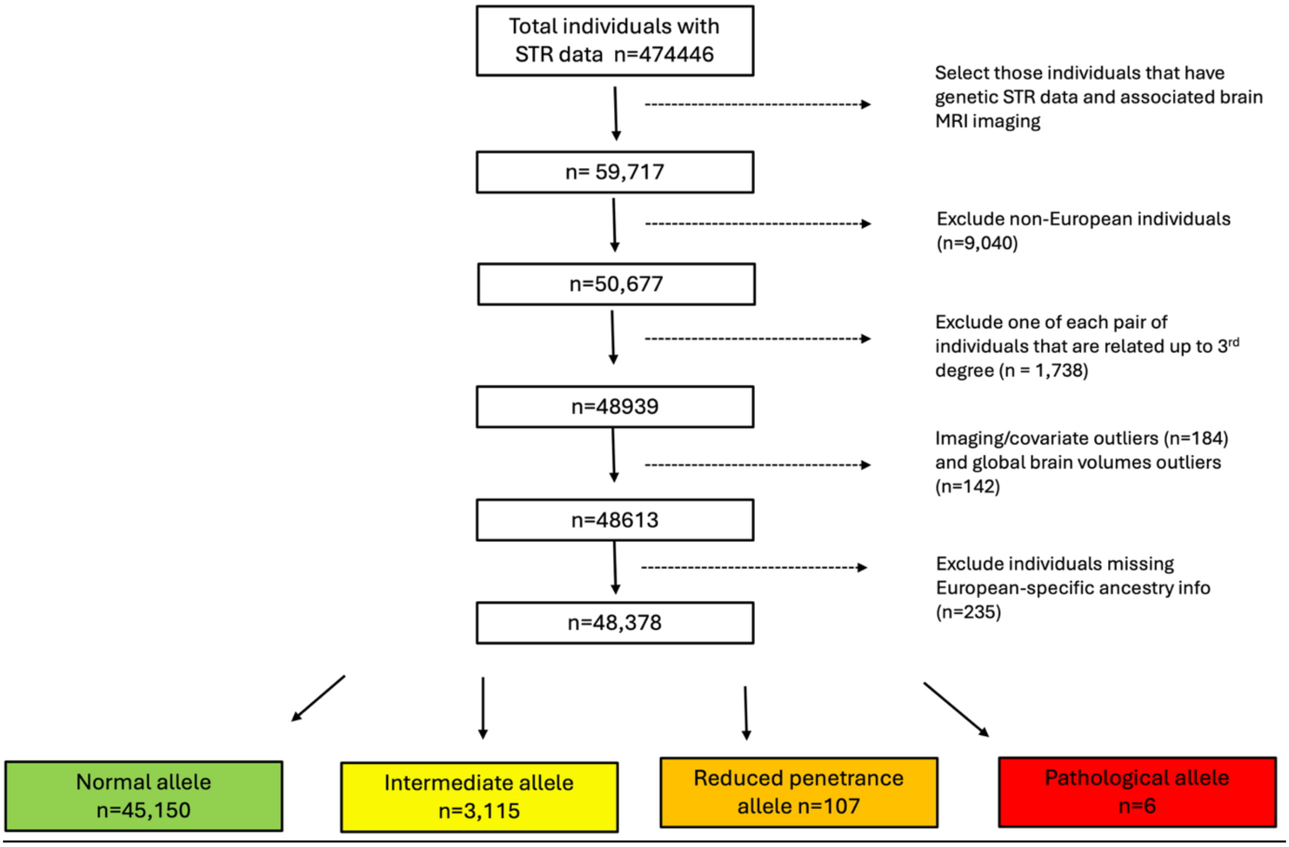

**Supplementary Figure SF2:** *HTT* CAG repeat length versus residualised subcortical volumes (UK Biobank). Panels A–D: amygdala, accumbens, caudate, pallidum. Points show mean residuals by CAG bin (midpoints ±SE), coloured by allele range (≤26, 27–35, ≥36). Dashed line connects bins ≤35; solid grey line (±95% CI) shows the linear fit within ≤35, extrapolated. Tables report M1: CAG main effect; M2: deviation for reduced penetrance and pathogenic alleles from extrapolated normal and intermediate range trend; M3: age × CAG interaction.

#### A. Amygdala

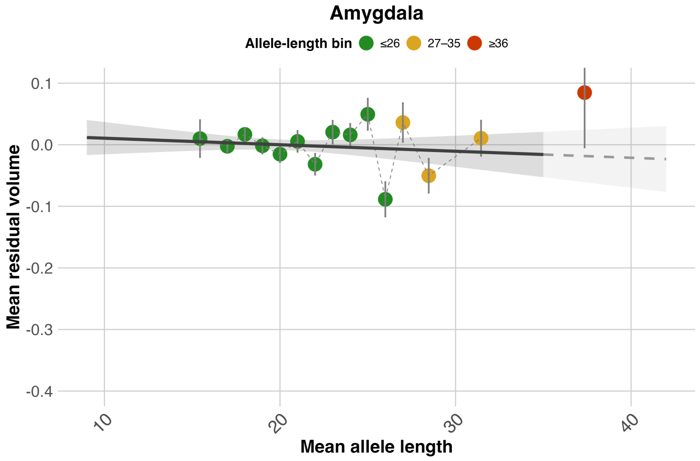


**Summary metrics (M1–M3)**

| Metric | Estimate | SE | p_value | N | CAG_range |
|---|---|---|---|---|---|
| M1 (CAG effect) | -0.003693 | 0.004322 | 0.392795 | 44,032 | ≤35 |
| M2 (path deviation) | 0.103000 | 0.089010 | 0.249691 | 105 | ≥36 vs ≤35 |
| M3 (age×CAG) | -0.420000 | 0.908100 | 0.643702 | 44,032 | ≤35 |

#### B. Accumbens

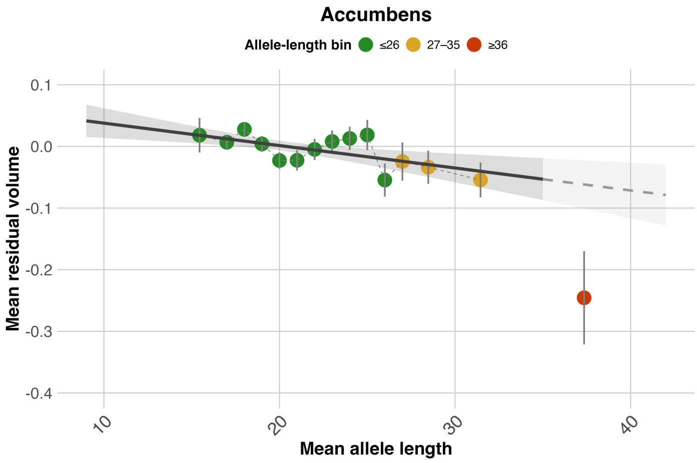


**Summary metrics (M1–M3)**

| Metric | Estimate | SE | p_value | N | CAG_range |
|---|---|---|---|---|---|
| M1 (CAG effect) | -0.0127 | 0.003977 | 0.001411 | 44,049 | ≤35 |
| M2 (path deviation) | -0.1839 | 0.074120 | 0.014706 | 105 | ≥36 vs ≤35 |
| M3 (age×CAG) | -2.9630 | 0.835700 | 3.93e-04 | 44,049 | ≤35 |

#### C. Caudate

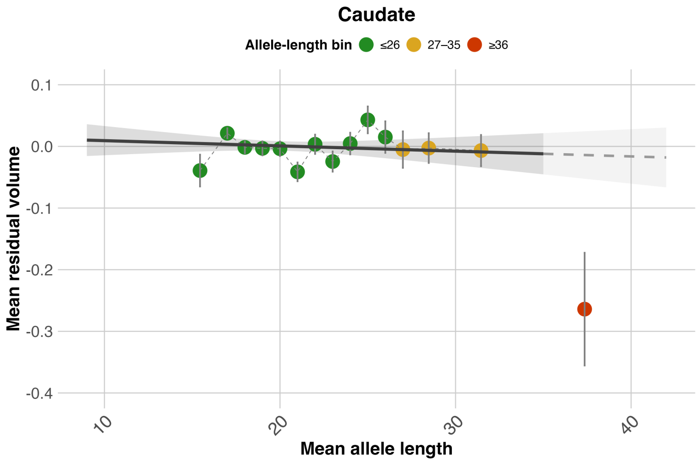


**Summary metrics (M1–M3)**

| Metric | Estimate | SE | p_value | N | CAG_range |
|---|---|---|---|---|---|
| M1 (CAG effect) | -0.002982 | 0.003911 | 0.445772 | 44,031 | ≤35 |
| M2 (path deviation) | -0.250000 | 0.091370 | 0.007311 | 105 | ≥36 vs ≤35 |
| M3 (age×CAG) | 0.097250 | 0.821800 | 0.905800 | 44,031 | ≤35 |

#### D. Pallidum

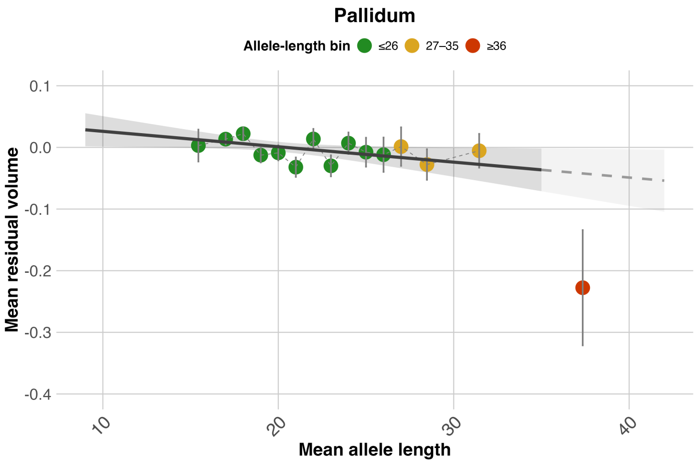


**Summary metrics (M1–M3)**

| Metric | Estimate | SE | p_value | N | CAG_range |
|---|---|---|---|---|---|
| M1 (CAG effect) | -0.008734 | 0.004065 | 0.031679 | 43,994 | ≤35 |
| M2 (path deviation) | -0.185400 | 0.093390 | 0.049804 | 105 | ≥36 vs ≤35 |
| M3 (age×CAG) | -0.352600 | 0.853700 | 0.679605 | 43,994 | ≤35 |

**Supplementary Figure SF3:** *HTT* CAG repeat length versus residualised subcortical volumes (UK Biobank). Panels E–G: putamen, thalamus, hippocampus. Points show mean residuals by CAG bin (midpoints ±SE), coloured by allele range (≤26, 27–35, ≥36). Dashed line connects bins ≤35; solid grey line (±95% CI) shows the linear fit within ≤35, extrapolated. Tables report M1: CAG main effect; M2: deviation for reduced penetrance and pathogenic alleles from extrapolated normal and intermediate range trend; M3: age × CAG interaction.

### E. Putamen

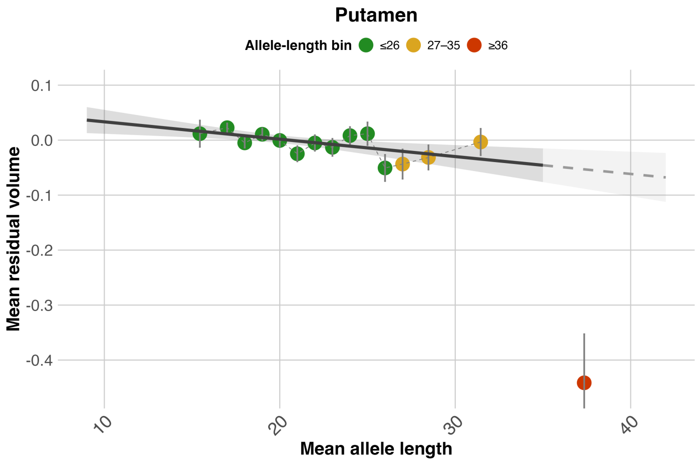


**Summary metrics (M1–M3)**

| Metric | Estimate | SE | p_value | N | CAG_range |
|---|---|---|---|---|---|
| M1 (CAG effect) | -0.01105 | 0.003585 | 0.002060 | 44,028 | ≤35 |
| M2 (path deviation) | -0.38830 | 0.088190 | 2.6e-05 | 105 | ≥36 vs ≤35 |
| M3 (age×CAG) | -0.60870 | 0.753000 | 0.418928 | 44,028 | ≤35 |

### F. Thalamus

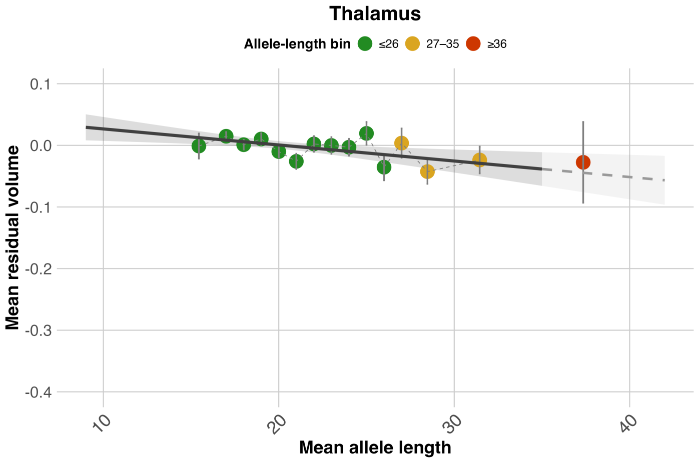


**Summary metrics (M1–M3)**

| Metric | Estimate | SE | p_value | N | CAG_range |
|---|---|---|---|---|---|
| M1 (CAG effect) | -0.009092 | 0.003199 | 0.004476 | 44,016 | ≤35 |
| M2 (path deviation) | 0.016970 | 0.065430 | 0.795857 | 105 | ≥36 vs ≤35 |
| M3 (age×CAG) | -0.292200 | 0.671900 | 0.663678 | 44,016 | ≤35 |

### G. Hippocampus

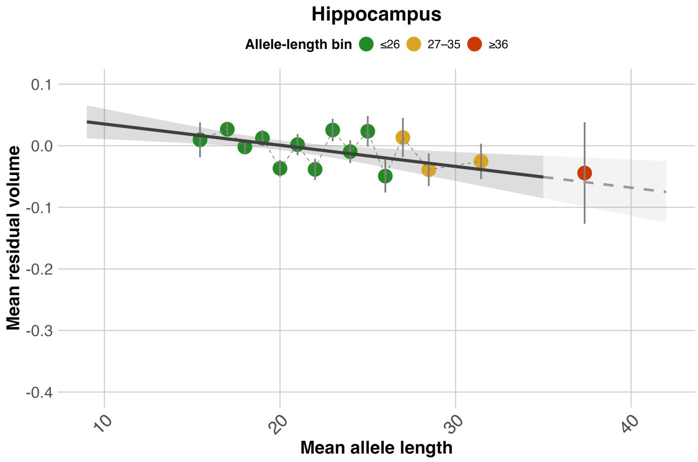


**Summary metrics (M1–M3)**

| Metric | Estimate | SE | p_value | N | CAG_range |
|---|---|---|---|---|---|
| M1 (CAG effect) | -0.01203 | 0.004042 | 0.002914 | 44,033 | ≤35 |
| M2 (path deviation) | 0.01465 | 0.080920 | 0.856695 | 105 | ≥36 vs ≤35 |
| M3 (age×CAG) | -1.76400 | 0.849100 | 0.037742 | 44,033 | ≤35 |

**Supplementary Figure SF4:** *HTT* CAG repeat length versus residualised global brain volumes (UK Biobank). Panels H–K: ventricles, CSF, subcortical grey matter, cortical grey matter. Points show mean residuals by CAG bin (midpoints ±SE), coloured by allele range (≤26, 27–35, ≥36). Dashed line connects bins ≤35; solid grey line (±95% CI) shows the linear fit within ≤35, extrapolated. Tables report M1: CAG main effect; M2: deviation for reduced penetrance and pathogenic alleles from extrapolated normal and intermediate range trend; M3: age × CAG interaction.

### H. Ventricles

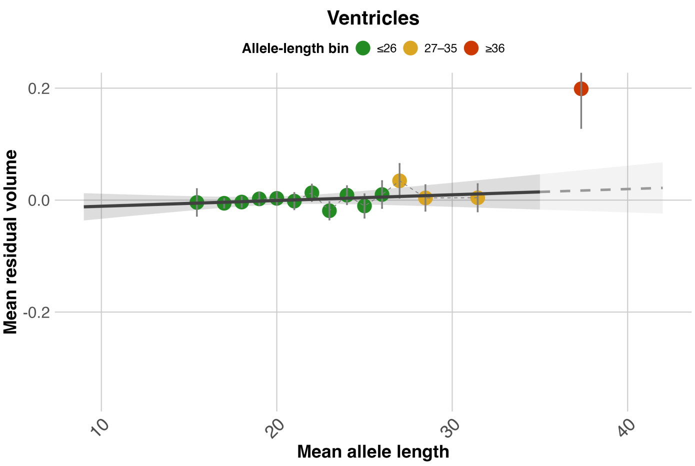


**Summary metrics (M1–M3)**

| Metric | Estimate | SE | p_value | N | CAG_range |
|---|---|---|---|---|---|
| M1 (CAG effect) | 0.003582 | 0.003692 | 0.331945 | 44,058 | ≤35 |
| M2 (path deviation) | 0.181800 | 0.069990 | 0.010753 | 105 | ≥36 vs ≤35 |
| M3 (age×CAG) | 0.739800 | 0.776000 | 0.340386 | 44,058 | ≤35 |

### I. CSF

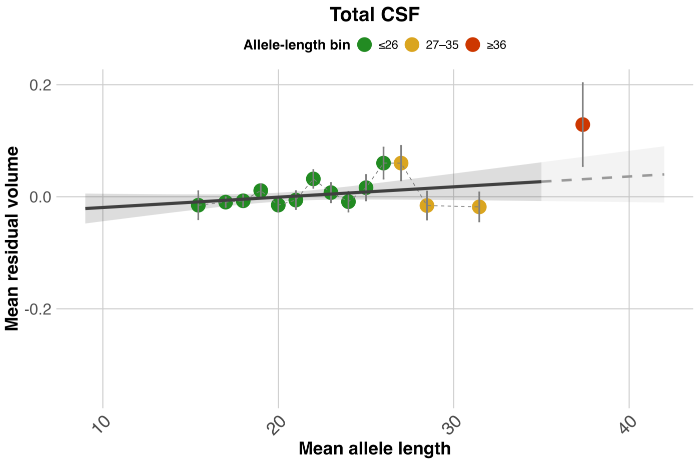


**Summary metrics (M1–M3)**

| Metric | Estimate | SE | p_value | N | CAG_range |
|---|---|---|---|---|---|
| M1 (CAG effect) | 0.00644 | 0.004048 | 0.111609 | 43,246 | ≤35 |
| M2 (path deviation) | 0.09752 | 0.074060 | 0.190851 | 104 | ≥36 vs ≤35 |
| M3 (age×CAG) | 0.20440 | 0.844000 | 0.808629 | 43,246 | ≤35 |

### J. Subcortical Grey Matter

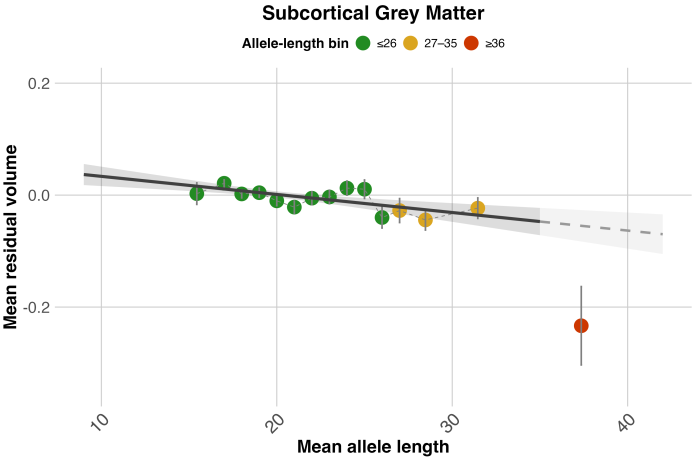


**Summary metrics (M1–M3)**

| Metric | Estimate | SE | p_value | N | CAG_range |
|---|---|---|---|---|---|
| M1 (CAG effect) | -0.01127 | 0.002858 | 8.03e-05 | 43,246 | ≤35 |
| M2 (path deviation) | -0.17850 | 0.069800 | 0.011981 | 104 | ≥36 vs ≤35 |
| M3 (age×CAG) | -0.57150 | 0.596000 | 0.337626 | 43,246 | ≤35 |

### K. Cortical Grey matter

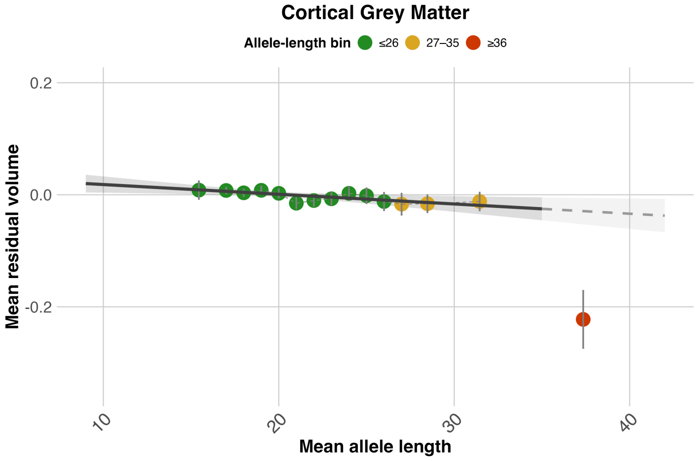


**Summary metrics (M1–M3)**

| Metric | Estimate | SE | p_value | N | CAG_range |
|---|---|---|---|---|---|
| M1 (CAG effect) | -0.006058 | 0.002394 | 0.011398 | 44,058 | ≤35 |
| M2 (path deviation) | -0.193300 | 0.050890 | 2.46e-04 | 105 | ≥36 vs ≤35 |
| M3 (age×CAG) | -0.114700 | 0.503200 | 0.819691 | 44,058 | ≤35 |

**Supplementary Figure SF5:** *HTT* CAG repeat length versus residualised global brain volumes (UK Biobank). Panels L–N: total grey matter, total white matter, total grey+white matter. Points show mean residuals by CAG bin (midpoints ±SE), coloured by allele range (≤26, 27–35, ≥36). Dashed line connects bins ≤35; solid grey line (±95% CI) shows the linear fit within ≤35, extrapolated. Tables report M1: CAG main effect; M2: deviation for reduced penetrance and pathogenic alleles from extrapolated normal and intermediate range trend; M3: age × CAG interaction.

#### L. Total Grey Matter

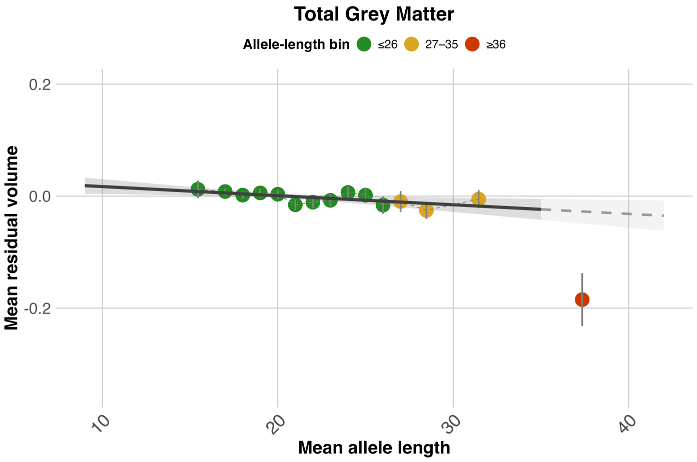


**Summary metrics (M1–M3)**

| Metric | Estimate | SE | p_value | N | CAG_range |
|---|---|---|---|---|---|
| M1 (CAG effect) | -0.005681 | 0.002187 | 0.009400 | 44,058 | ≤35 |
| M2 (path deviation) | -0.157500 | 0.045670 | 8.14e-04 | 105 | ≥36 vs ≤35 |
| M3 (age×CAG) | -0.360300 | 0.459700 | 0.433286 | 44,058 | ≤35 |

#### M. Total White Matter

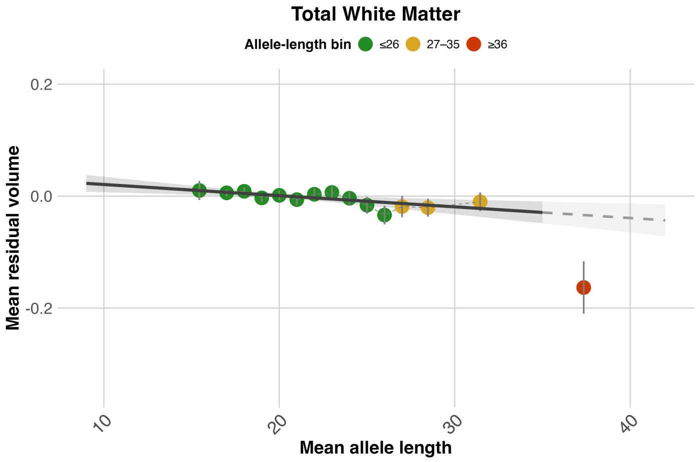


**Summary metrics (M1–M3)**

| Metric | Estimate | SE | p_value | N | CAG_range |
|---|---|---|---|---|---|
| M1 (CAG effect) | -0.006985 | 0.002282 | 0.002213 | 44,058 | ≤35 |
| M2 (path deviation) | -0.129500 | 0.045070 | 0.004938 | 105 | ≥36 vs ≤35 |
| M3 (age×CAG) | -0.272900 | 0.479700 | 0.569489 | 44,058 | ≤35 |

#### N. Total Grey and White Matter

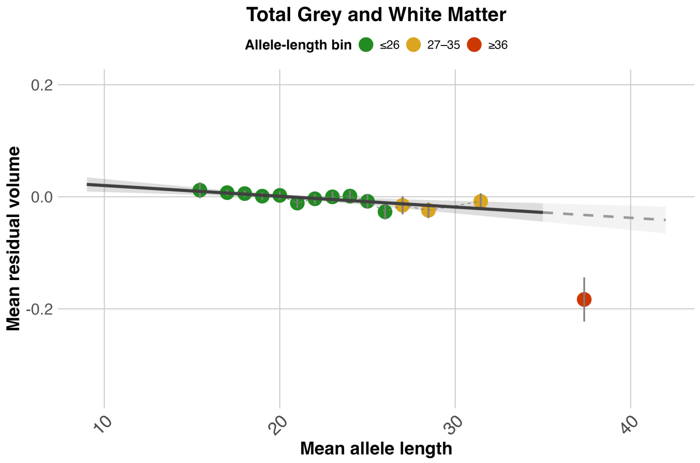


**Summary metrics (M1–M3)**

| Metric | Estimate | SE | p_value | N | CAG_range |
|---|---|---|---|---|---|
| M1 (CAG effect) | -0.006721 | 0.001925 | 4.8e-04 | 44,058 | ≤35 |
| M2 (path deviation) | -0.150800 | 0.037870 | 1.27e-04 | 105 | ≥36 vs ≤35 |
| M3 (age×CAG) | -0.332100 | 0.404500 | 0.411628 | 44,058 | ≤35 |

**Supplementary Figure SF6**: Age-stratified association between *HTT* CAG repeat length and residualised subcortical volumes. Points show mean residuals by CAG bin (midpoints ±SE), coloured by allele range (≤26, 27–35). Solid lines show linear fits for ≤66 years (dark grey) and ≥67 years (maroon), restricted to CAG ≤35.

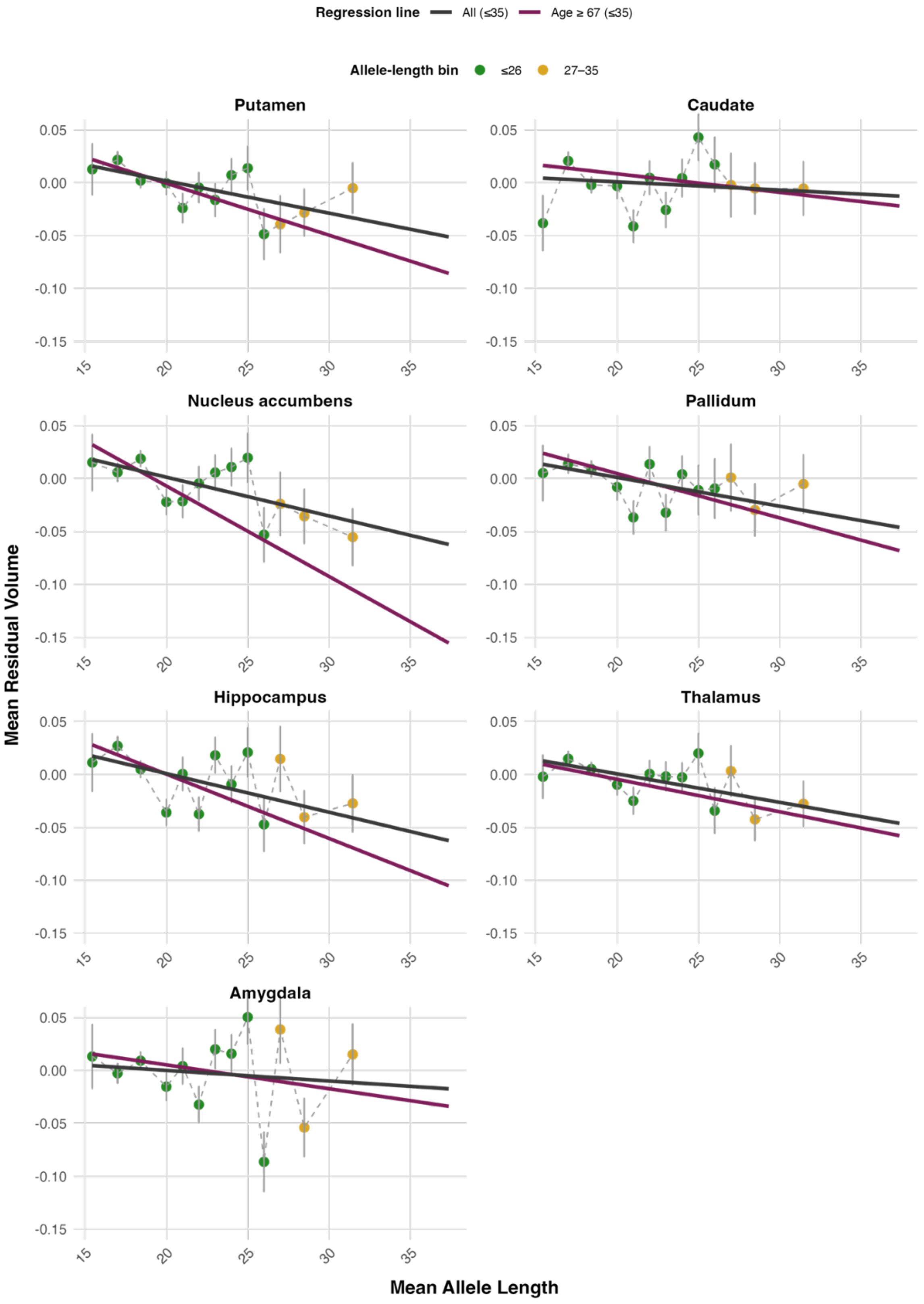

**Supplementary Figure SF7:** Age-stratified association between *HTT* CAG repeat length and residualised global volumes. Points show mean residuals by CAG bin (midpoints ±SE), coloured by allele range (≤26, 27–35). Solid lines show linear fits for ≤66 years (dark grey) and ≥67 years (maroon), restricted to CAG ≤35.

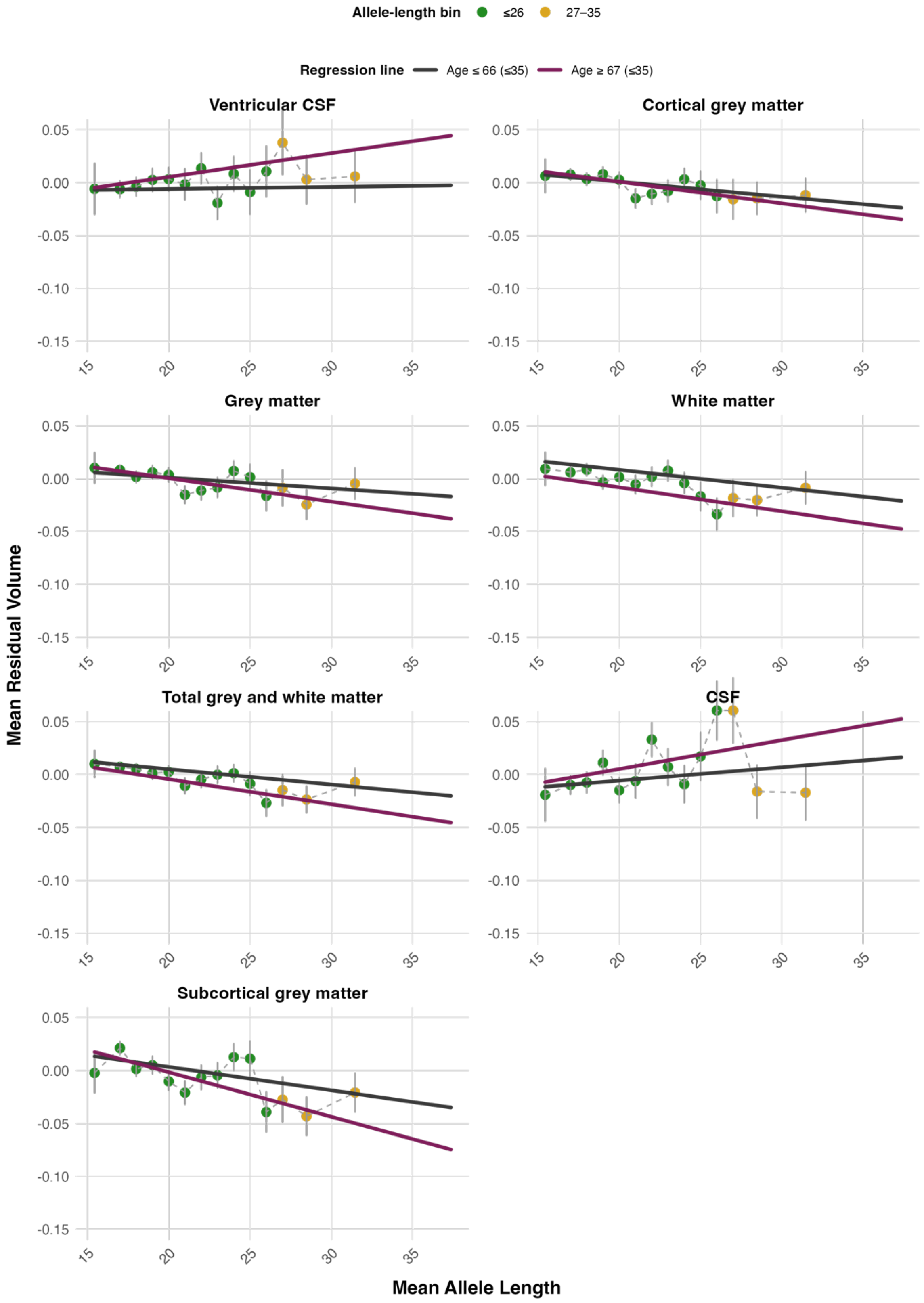

**Supplementary Figure SF8**: Kaplan–Meier curves showing age-dependent dementia-free survival (A) and anxiety-free survival (B) across three *HTT* CAG-repeat length categories: normal allele carriers (CAG length ≤ 26, green), intermediate allele carriers (CAG length 27–35, yellow), and reduced-penetrance or pathogenic alleles (CAG length ≥36, red).

A

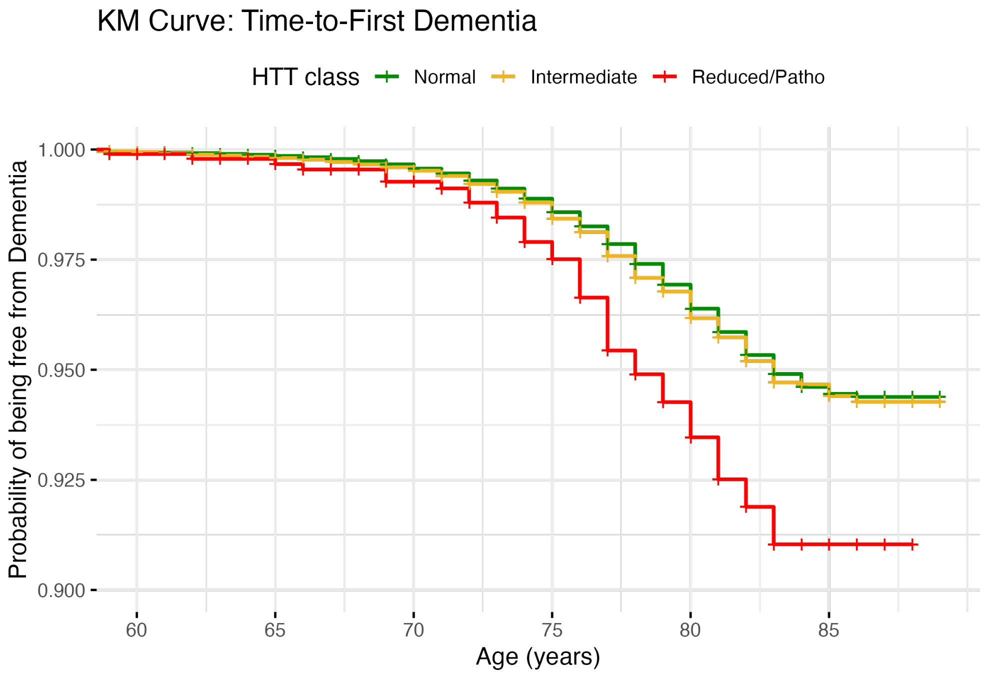


B

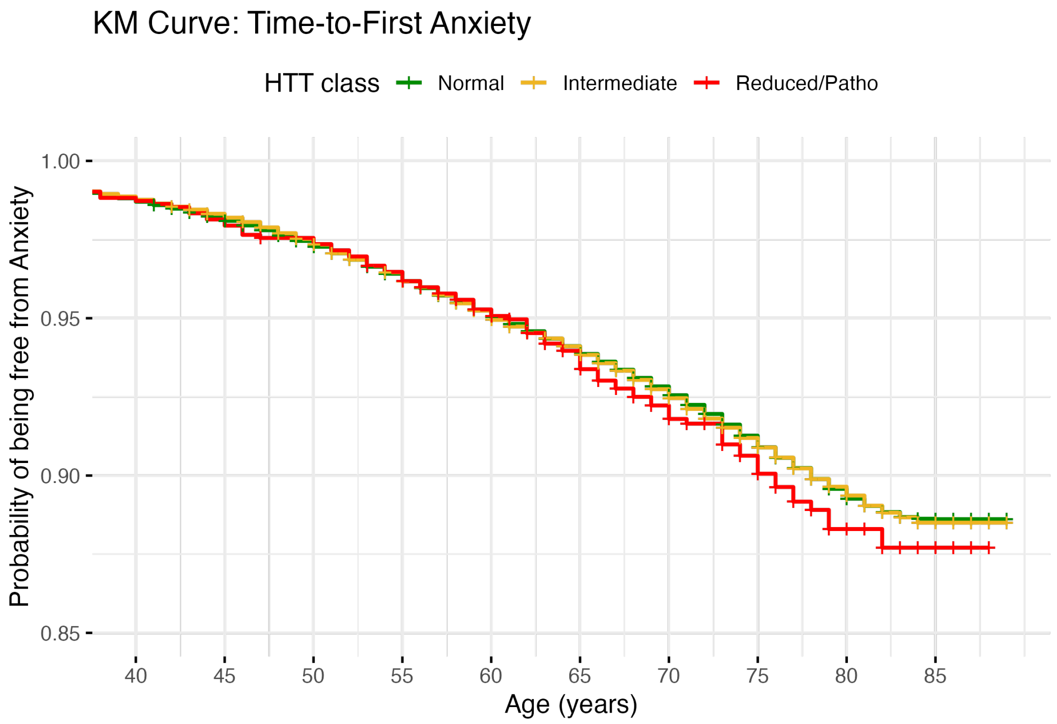

**Supplementary Figure SF9:** Residualised cognitive phenotypes vs *HTT* CAG repeat length in UK Biobank. Panels A–C show Reaction Time, Fluid Intelligence, and Symbol Digit Substitution. Points denote mean residuals by CAG bin (±SE) from covariate-only models (age modelled as a second-degree orthogonal polynomial, sex, age×sex, and the first ten ancestry principal components). Grey line (±95% CI) shows the linear trend fitted within CAG ≤35 and extrapolated beyond. Tables report M1: CAG main effect; M2: deviation for reduced penetrance and pathogenic alleles from extrapolated normal and intermediate range trend.

### A. Reaction Time

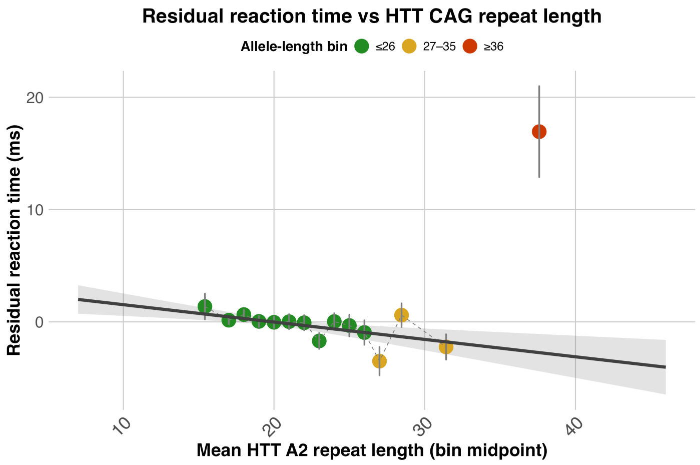


**Summary metrics (M1–M2)**

| Metric | Estimate | SE | p_value | N | CAG_range |
|---|---|---|---|---|---|
| M1 (CAG effect) | -0.1553 | 0.04764 | 0.001114 | 463,375 | ≤35 |
| M2 (path deviation) | 19.6800 | 4.04300 | 1.32e-06 | 1,009 | ≥36 vs ≤35 |

### B. Fluid Intelligence

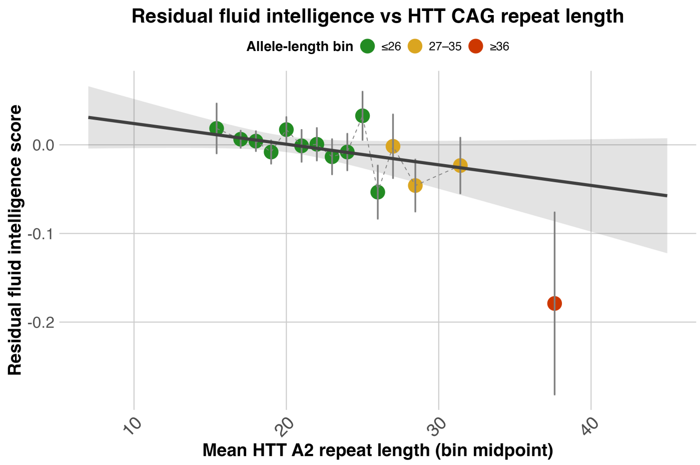


**Summary metrics (M1–M2)**

| Metric | Estimate | SE | p_value | N | CAG_range |
|---|---|---|---|---|---|
| M1 (CAG effect) | -0.002343 | 0.001322 | 0.076328 | 205837 | ≤35 |
| M2 (path deviation) | -0.138700 | 0.102600 | 0.177221 | 440 | ≥36 vs ≤35 |

### C. Symbol Digit Substitution

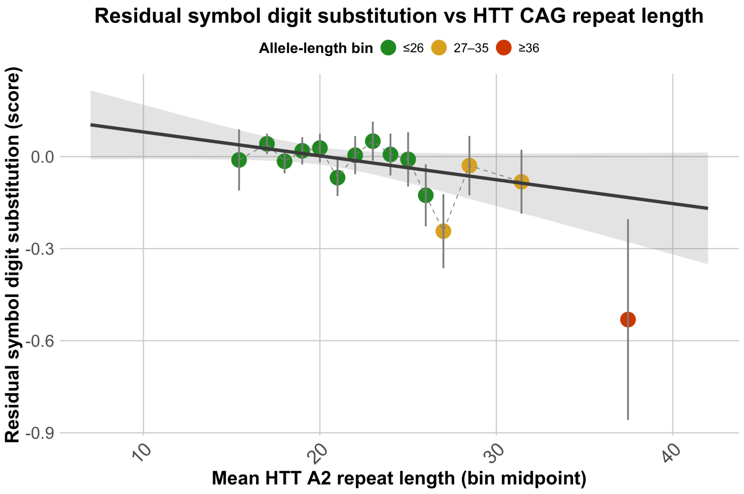


**Summary metrics (M1–M2)**

| Metric | Estimate | SE | p_value | N | CAG_range |
|---|---|---|---|---|---|
| M1 (CAG effect) | -0.007795 | 0.004206 | 0.063856 | 104,621 | ≤35 |
| M2 (path deviation) | -0.397600 | 0.324300 | 0.221564 | 210 | ≥36 vs ≤35 |

**Supplementary Figure SF10:** Normative deviation in reaction time (A), fluid intelligence (B), and Symbol Digit Substitution (C) among carriers of pathogenic *HTT* CAG repeat expansions. The upper panel shows the reference distribution of z-scores in individuals with normal *HTT* alleles (A2 ≤ 26). Vertical ticks mark carriers of pathogenic alleles (CAG ≥ 40), coloured by repeat-length category (40–41, 42–43, 44–46). The lower panel shows these individuals by age at assessment. Vertical lines denote the mean (0 SD) and thresholds at 1, 1.645, and 2 SD. The x-axis is shared across panels.

A

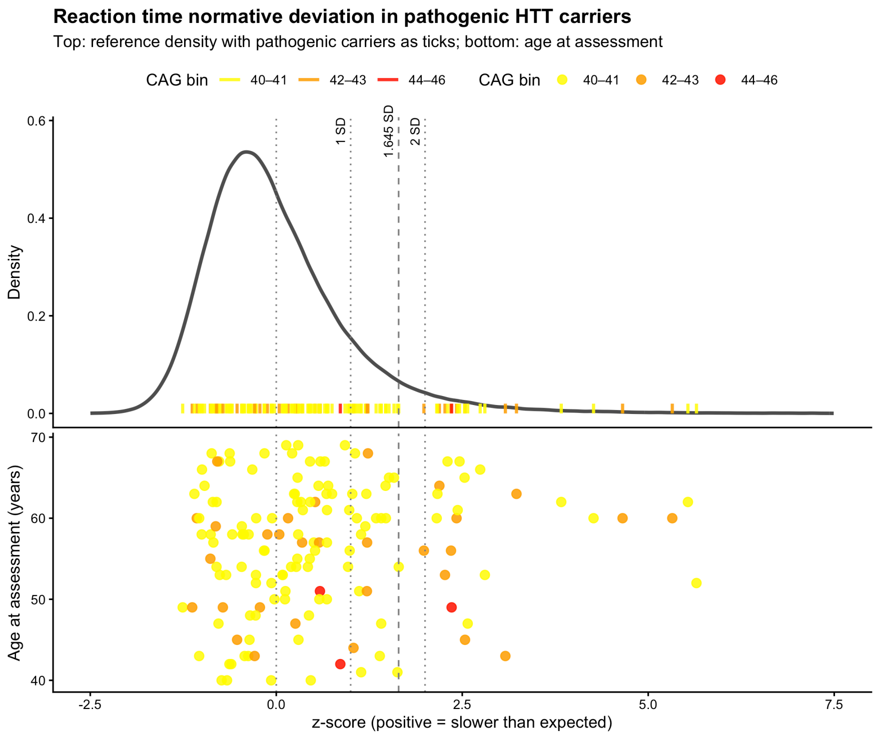

B

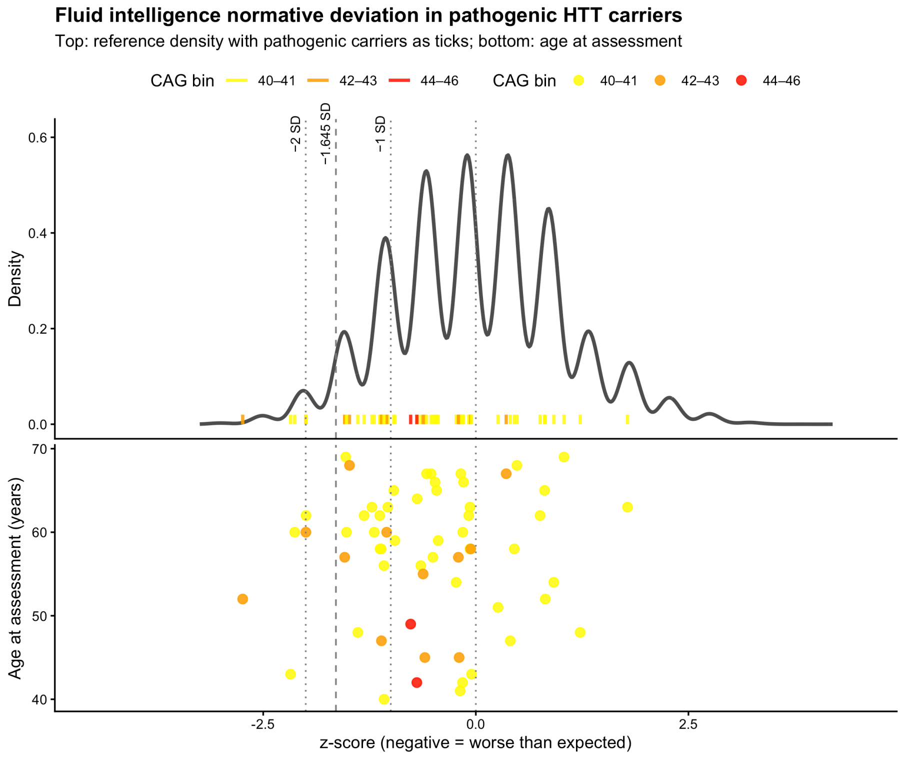
Fluid intelligence normative deviation in pathogenic HTT carriers
Top: reference density with pathogenic carriers as ticks; bottom: age at assessment
CAG bin 40–41 42–43 44–46 CAG bin 40–41 42–43 44–46
−2 SD
−1.645 SD
−1 SD
Density
0.6
0.4
0.2
0.0
Age at assessment (years)
70
60
50
40
-2.5
0.0
2.5
z-score (negative = worse than expected)

C

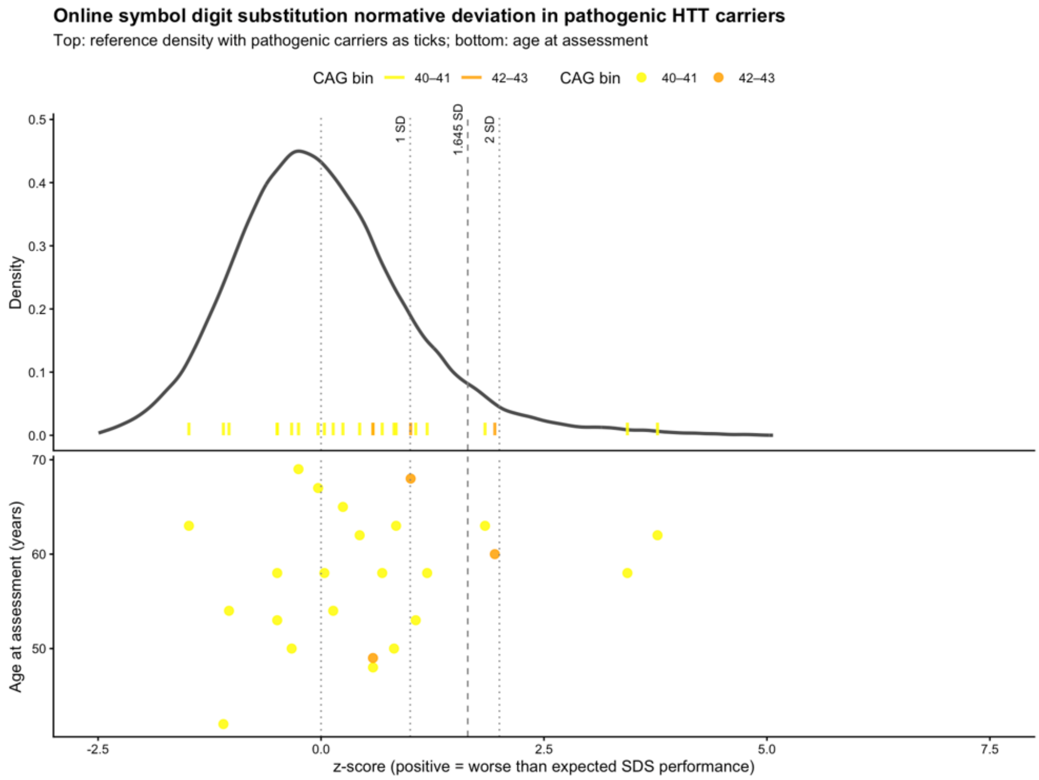
Online symbol digit substitution normative deviation in pathogenic HTT carriers
Top: reference density with pathogenic carriers as ticks; bottom: age at assessment
CAG bin 40–41 42–43 CAG bin 40–41 42–43
1 SD
1.645 SD
2 SD
Density
0.5
0.4
0.3
0.2
0.1
0.0
Age at assessment (years)
70
60
50
-2.5
0.0
2.5
5.0
7.5
z-score (positive = worse than expected SDS performance)

#### Supplementary Table ST1: Effect sizes for psychiatric phenotypes in full mixed ancestry and European cohorts

| Outcome | Contrast | HR | CI_low | CI_high | p | p_FDR_BH |
|---|---|---|---|---|---|---|
| Depression | Intermediate vs Normal | 1.054486 | 1.014958 | 1.095554 | 0.00649635 | 0.0259854 |
| Depression | Reduced/path vs Normal | 1.220459 | 1.0121549 | 1.471632 | 0.03693382 | 0.07386764 |
| Dementia | Intermediate vs Normal | 1.056566 | 0.9705264 | 1.150233 | 0.20420869 | 0.29053532 |
| Dementia | Reduced/path vs Normal | 1.809133 | 1.297983 | 2.521575 | 0.00046613 | 0.00372907 |
| Anxiety | Intermediate vs Normal | 1.00674 | 0.966941 | 1.048177 | 0.74412074 | 0.74412074 |
| Anxiety | Reduced/path vs Normal | 1.084484 | 0.8867008 | 1.326383 | 0.42983812 | 0.49124357 |
| Delirium | Intermediate vs Normal | 1.055197 | 0.9687607 | 1.149345 | 0.21790149 | 0.29053532 |
| Delirium | Reduced/path vs Normal | 1.547386 | 1.0811569 | 2.214668 | 0.01700819 | 0.04535518 |

#### Supplementary Table ST2: Sample sizes and case counts for psychiatric and clinical outcomes

| Phenotype | Definition / Source | Total N | Cases (N) | Notes |
|---|---|---|---|---|
| Depression | MHQ (MHQ1/MHQ2 self-report) | 201,987 | 43,000 | Mental Health questionnaire-based phenotype |
| Anxiety | ICD-10 F41 | 469,196 | 39,788 | Case status derived from linked ICD-10 records. |
| Delirium | ICD-10 F05 | 469,202 | 8,529 | Case status derived from linked ICD-10 records. |
| Dementia | ICD-10 diagnoses (all-cause dementia: F00, F01, F02 and F03) | 469,198 | 8,645 | Case status derived from linked ICD-10 records. |

#### Supplementary Table ST3: *HTT* CAG Repeat Length and Cognitive Measures

| Phenotype | Anc | M1 β | M1 SE | M1 p-value | M1 N | M2 Δ | M2 SE | M2 p-value | M2 N | M1 $p_{FDR}$ | M2 $p_{FDR}$ |
|---|---|---|---|---|---|---|---|---|---|---|---|
| Reaction time | All | -0.155 | 0.0476 | 0.0011 | 463375 | 19.7 | 4.04 | 1.32e-6 | 1009 | 0.0067 | 7.90e-6 |
| Reaction time | EUR | -0.150 | 0.0547 | 0.0060 | 325691 | 21.2 | 4.70 | 7.70e-6 | 720 | 0.0181 | 2.31e-5 |
| Fluid intelligence | All | -0.00234 | 0.0013 | 0.0763 | 205837 | -0.139 | 0.103 | 0.177 | 440 | 0.0961 | 0.213 |
| Fluid intelligence | EUR | -0.00376 | 0.0015 | 0.0166 | 145661 | -0.101 | 0.124 | 0.419 | 322 | 0.0331 | 0.419 |
| Symbol digit substitution | All | -0.00669 | 0.0040 | 0.0961 | 114929 | -0.433 | 0.310 | 0.165 | 230 | 0.0961 | 0.213 |
| Symbol digit substitution | EUR | -0.00804 | 0.0048 | 0.0951 | 78187 | -0.589 | 0.365 | 0.109 | 165 | 0.0961 | 0.213 |
